# A phenotype-to-mechanism framework links phenome-wide comorbidity architecture to molecular mechanisms and therapeutic discovery in complex diseases

**DOI:** 10.64898/2026.05.13.26353128

**Authors:** Wei-Ting Wang, Manqi Zhou, Jie Tong, Meng-Ju Lin, Alison Ke, Meihan Wei, Zhenxing Xu, Hansen Tai, Aarthi Parvathaneni, Khyla T. Hill, Steven R. Cohen, Lynn Petukhova, Ernest S. Chiu, Fei Wang, Catherine P. Lu, Chang Su

## Abstract

Complex human diseases exhibit substantial clinical heterogeneity driven by poorly understood molecular mechanisms, while many also lack sufficient molecular and omics data for mechanistic investigation, hindering therapeutic development. We introduce PiMInfer, a “phenotype-to-mechanism” framework that leveraged largely available real-world clinical data-based deep phenotypic characterizations with a biomedical knowledge graph approach to resolve disease clinical heterogeneity into phenotype-informed molecular modules, thereby accelerating therapeutic target discovery. We applied PiMInfer to investigate Hidradenitis Suppurativa (HS), an autoimmune skin disease with poorly understood pathogenesis and limited treatment options. PiMInfer identified a coherent, phenotype-informed HS gene module (PiHSM) and functional endotypes, which were validated using multimodal evidence. *In silico* drug repurposing using PiHSM prioritized Carfilzomib, targeting the immunoproteasome subunit *PSMB9*, essential for MHC Class I antigen presentation. Preclinical testing using human patient lesional skin explants confirmed its anti-inflammatory activity and demonstrated a significant downregulation of IFN-γ, IL-17, and mTOR signaling pathways within HS lesional microenvironment through single-cell RNA sequencing. PiHSM-based network predictions further suggest a potential enhanced efficacy of combining Carfilzomib with approved HS agents. Collectively, PiMInfer provides a scalable framework that bridges real-world phenome-wide comorbid associations to mechanism-anchored therapeutic discovery, enabling a paradigm shift in precision medicine approaches for complex diseases with limited molecular characterization and in need of better therapeutic strategies.

## Introduction

Translational research for human diseases with poorly understood pathogenesis and complex comorbidity patterns faces critical challenges - complex diseases exhibit high clinical heterogeneity as results of tangled networks of interacting factors and pathways with patient-to-patient variabilities. The lack of robust experimental models and predictive biomarkers severely impedes our capacity to investigate molecular mechanisms, perform pre-clinical trials, and to measure therapeutic efficacy. While large-scale genetics and other omics datasets have become central to disease mechanism discovery, their translational utility is often restricted by the absence of deep phenotypic characterization needed to connect molecular findings with real-world clinical heterogeneity, as well as by the limited availability of high-quality omics data for many rare or understudied diseases. Together, these barriers create a persistent gap in the translational pipelines, preventing the conversion of experimental discoveries into real-world patient benefits^1,2^.

The past decades’ widespread adoption of clinical data infrastructures has produced massive volumes of real-world data (RWD), most notably electronic health records (EHRs). EHRs capture longitudinal, patient-level information, including historical diagnoses, medication, laboratory results, procedures, and outcomes, reflecting real-world clinical practice. These resources enable the efficient construction of richly phenotyped disease cohorts to generate real-world evidence^3^. However, because EHRs primarily capture bedside clinical manifestations rather than underlying molecular mechanisms, there remains a pressing need of translating these phenotypic signals into biological insights.

On the other hand, biomedical knowledge graphs (BKGs) are heterogeneous networks that assembles a vast and continually expanding set of biomedical entities across modalities including phenotypes (diseases, symptoms), genes, pathways, drugs, etc. and the diverse relations among them, all supported by existing experimental and clinical evidence^4^. We have previously developed a comprehensive BKG, the integrative biomedical knowledge hub (iBKH), through harmonizing diverse biomedical knowledge sources into a unified framework^5^. Furthermore, because BKG encodes the complex, interconnected architecture of human biology and diseases, it inherently aligns with the principle of network medicine^4,6^ and potentially provides a feasible scaffold for mapping phenotypic signals onto disease mechanisms. Prior studies have shown that incorporating BKGs into network medicine workflows enables the propagation of EHR-derived phenotypic signals across modalities^7–9^; however, these efforts have not extended to constructing disease-specific molecular networks, performing experimental validation, or advancing downstream therapeutic development.

Leveraging the real-world phenotypic architecture derived from large-scale EHRs—particularly the full spectrum of comorbid conditions associated with a given disease— and coupling them with BKG-driven mechanistic inference, we developed PiMInfer (phenotype-informed disease mechanism inference), a generalizable framework for disease-associated gene module identification and *in silico* drug repurposing (**Fig. 1**). In this study, we applied the PiMInfer framework to study Hidradenitis Suppurativa (HS), a chronic, painful, recurrent inflammatory skin disease with substantial clinical heterogeneity and comorbidity burden, poorly understood pathogenesis, and limited therapeutic options^10,11^. Using two large-scale EHR repositories in combination with iBKH^5^, we constructed a coherent phenotype-informed gene module for HS (PiHSM) comprising distinct functional endotypes. These PiHSM module genes were validated using multimodal biological evidence from human patient skin tissues, confirming their relevance to HS pathophysiology. Furthermore, we demonstrated that PiHSM enables robust prediction of therapeutic targets for *in silico* drug repurposing, and validated by identifying Carfilzomib, a Food and Drug Administration (FDA)–approved drug, as a promising therapeutic candidate for HS. PiHSM-based network analyses further suggested potential enhanced efficacy when Carfilzomib is combined with approved TNF-α and IL-17 blockades for HS. Together, this PiMInfer framework provides a scalable route from real-world phenotypic evidence to mechanism-guided therapeutic insights, leveraging large and rapidly growing EHRs to accelerate drug discovery—especially for understudied diseases with high complexity.

**Figure 1.**
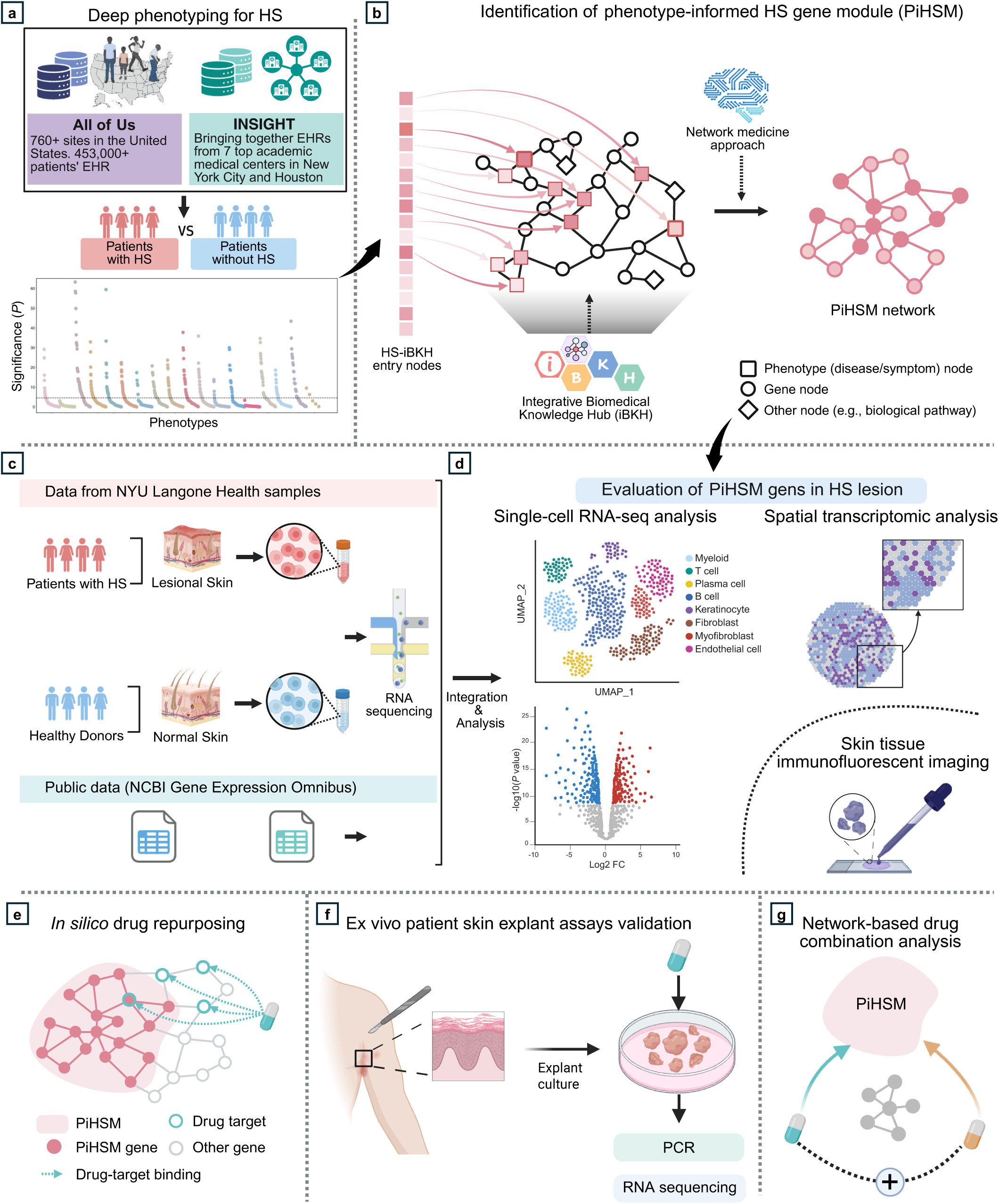
Overview of study pipeline. We demonstrated our PiMInfer framework through study of Hidradenitis Suppurativa (HS). (**a**) Deep phenotyping over two large-scale electronic health record (EHR) data repositories, All of Us and INSIGHT, systematically identified and validated robust comorbidity profiles of HS. (**b**) We next injected the phenotypic signals (i.e., comorbidity profiles) into our biomedical knowledge graph (BKG), i.e., iBKH, and applied a network-based approach to propagate the signals from phenome to genome through iBKH network structure. This resulted in the discovery of a coherent disease molecular network for HS, i.e., PiHSM. (**c-d**) We validated the PiHSM module genes using multimodal biological evidence from human patients, including single-cell and spatial transcriptomics and immunofluorescent microscopy. (**e**) *In silico* drug repurposing was performed based on the identified PiHSM using a network proximity-based approach. (**f**) We experimentally examined treatment efficacy of drug candidates through preclinical testing using human patient lesional skins. Specifically, we tested a drug’s anti-inflammatory activity within the HS lesion microenvironment by quantitative PCR and single-cell RNA sequencing. (**g**) Network-based analysis evaluating potential combination therapy with the FDA-approved HS treatments (Secukinumab and Adalimumab). Abbreviations: HS, Hidradenitis Suppurativa.

## Results

### Construction of comprehensive comorbidity architecture of Hidradenitis Suppurativa using large-scale real-world EHR

We used two large-scale EHR repositories, the All of Us (AoU) Research Program and the INSIGHT Clinical Research Network. From the AoU data, we identified 2,191 individuals with HS and 214,750 individuals without HS as the control group who met the eligibility criteria (**Extended Data Fig. 1**). From the INSIGHT data, we identified 9,479 individuals with HS and 308,125 individuals without HS who met the eligibility criteria (**Extended Data Fig. 1**). Demographic characteristics of the studied individuals are described in **Table 1**.

**Table 1.** Demographic characteristics of the studied population in the All of Us and INSIGHT data repositories.

| Characteristic | All of Us |  |  |  |  | INSIGHT |  |  |  |  |
| --- | --- | --- | --- | --- | --- | --- | --- | --- | --- | --- |
|  | Overall | HS Group | Control Group | Matched control group | SMD | Overall | HS Group | Control Group | Matched control group | SMD |
| # of patients | (N= 24,101) | (N = 2,191) | (N =214,751) | (N = 21,910) |  | (N= 317,604) | (N = 9,479) | (N =308,125) | (N =94,750) |  |
| Age, mean (SD), yr | 48.9 (15.3) | 48.6 (14.7) | 55.4 (16.7) | 48.9 (15.4) | -0.02 | 50.13 (18.9) | 43.28 (15.6) | 50.34 (18.9) | 43.21 (15.78) | 0.00 |
| Sex, N (%) |  |  |  |  |  |  |  |  |  |  |
| Female | 19,291 (80) | 1,727 (78.8) | 134,030 (62.4) | 17,564 (80.2) | -0.03 | 189,270 (59.6) | 7,012 (74) | 182,258 (59.2) | 68,402 (72.2) | 0.04 |
| Male | 4,626 (19.2) | 442 (20.2) | 78,410 (36.5) | 4,184 (19.1) | 0.03 | 128,271 (40.4) | 2,462 (26) | 125,809 (40.8) | 26,319 (27.8) | -0.04 |
| Race, N (%) |  |  |  |  |  |  |  |  |  |  |
| Black or African American | 9,633 (40) | 876 (40) | 39,453 (18.4) | 8,757 (40.0) | 0.00 | 39,494 (12.4) | 2,922 (30.8) | 36,572 (11.9) | 27,953 (29.5) | 0.03 |
| No answer race | 3,725 (15.5) | 359 (16.4) | 40,070 (18.7) | 3,366 (15.4) | 0.03 | 103,122 (32.5) | 3,318 (35) | 99,804 (32.4) | 34,598 (36.5) | -0.03 |
| Other race | 1,033 (4.3) | 111 (5.1) | 12,366 (1.7) | 922 (4.2) | 0.04 | 31,079 (9.8) | 646 (6.8) | 30,433 (9.9) | 6,226 (6.6) | 0.01 |
| White | 9,710 (40.3) | 845 (38.6) | 122,862 (57.2) | 8,865 (40.5) | -0.04 | 143,909 (45.3) | 2,593 (27.4) | 141,316 (45.9) | 25,973 (27.4) | 0.00 |
| Ethnicity, N (%) |  |  |  |  |  |  |  |  |  |  |
| Hispanic or Latino | 3,709 (15.4) | 359 (16.4) | 38,761 (18) | 3,350 (15.3) | 0.03 | 40,033 (12.6) | 1,850 (19.5) | 38,183 (12.4) | 19,351 (20.4) | -0.02 |
| Not Hispanic or Latino | 19,582 (81.2) | 1,745 (79.6) | 168,021 (78.2) | 17,837 (81.4) | -0.04 | 192,501 (60.6) | 5,571 (58.8) | 186,930 (60.7) | 54,560 (57.6) | 0.02 |
| No answer ethnicity | 508 (2.1) | 58 (2.6) | 5,791 (2.7) | 450 (2.1) | 0.04 | - | - | - | - | - |
| Other ethnicity | 302 (1.3) | 29 (1.3) | 2,178 (1) | 273 (1.2) | 0.01 | 85,070 (26.8) | 2,058 (21.7) | 83,012 (26.9) | 20,839 (22) | -0.01 |

Abbreviations: HS, Hidradenitis Suppurativa; SMD, standardized mean difference

We examined phenotypes at phenome wide, mapped to 1848 phecodes^12^ across 18 disease categories (**Methods**), and identified 448 comorbidities enriched in HS within the AoU cohort, which met the Bonferroni-corrected significance threshold (*P* < 0.05/1848 = 2.7×10^-5^) (**Fig. 2a-b** and **Supplementary Tables 1-2**). We observed strong associations between HS and other dermatological conditions, notably superficial cellulitis and abscess, cellulitis-related conditions affecting different body areas, carbuncle and furuncle, diseases of hair and hair follicles, and pilonidal cyst. Enrichment of systemic autoimmune disorders, including rheumatoid arthritis (RA), inflammatory bowel disease (IBD), and type 1 diabetes (T1D), aligns with the theories of autoimmune/autoinflammatory nature of HS^13–15^. Enrichment of obesity, type 2 diabetes (T2D), metabolic syndrome, polycystic ovarian syndrome, hyperlipidemia, hypertension, and tobacco smoking were supported by literature^16–19^; these conditions are important risk factors of cardiovascular diseases. Psychiatric disorders (e.g., depression, anxiety, etc.) and suicidality, substance use disorder, sleep problems, anemia, vitamin D & vitamin B-complex deficiency, and renal disorders were also enriched in HS and concordant with prior studies^20–22^.

**Figure 2.**
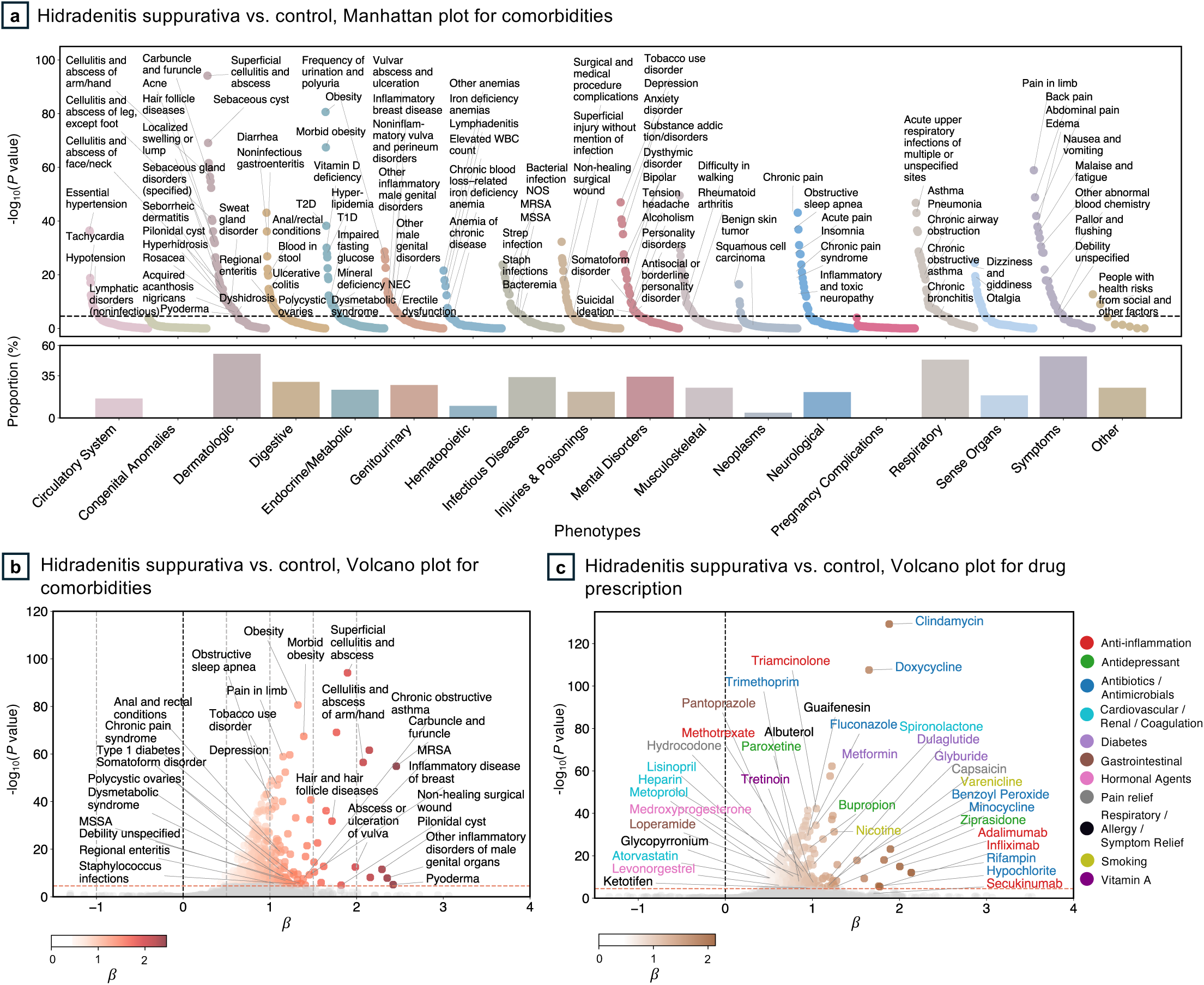
Association analyses identify comorbidity and medication enrichments in Hidradenitis Suppurativa (HS). (**a**) Manhattan plot (top) showing results of comorbidity association analysis. The dots represent phecodes, colored and grouped by phecode categories. The dashed horizontal line indicates Bonferroni-corrected significance threshold (*P* < 0.05/1848 = 2.7×10^-5^). The bar plot (bottom) illustrates the proportion of phecodes within each disease category significantly enriched in HS. (**b**) Volcano plot displaying estimated coefficients of the phenotypic associations. The dashed horizontal line indicates Bonferroni-corrected significance threshold. The dots represent phecodes, colored according to their estimated association coefficients. (**c**) Volcano plot showing results of medication association analysis. The dashed horizontal line indicates Bonferroni-corrected significance threshold. The dots represent medications, colored according to their estimated association coefficients. Abbreviations: HS, Hidradenitis Suppurativa; T1D, type 1 diabetes mellitus; T2D, type 2 diabetes mellitus; MRSA, methicillin-resistant Staphylococcus aureus; MSSA, methicillin-sensitive Staphylococcus aureus.

To assess reproducibility, we validated our findings by comparing HS patients with demographic-matched controls in the AoU, as well as with general and demographic-matched controls in the independent INSIGHT data (**Methods**). The associations derived in primary analysis remained reproducible (**Extended Data Fig. 2-3** and **Supplementary Tables 3-5**).

In addition, we examined associations between medication prescriptions with HS. Medications were mapped to 1,611 ingredients. A total of 238 medication prescriptions were enriched in HS within the AoU cohort, which met the Bonferroni-corrected significance threshold (*P* < 0.05/1611 = 3.1×10^-5^) (**Fig. 2c** and **Supplementary Table 6**). Notable medications include 1) TNF-α inhibitors and antineoplastic for HS treatments including adalimumab, infliximab (used to treat moderate-to-severe HS^23^ and methotrexate; 2) anti-inflammatory/anti-infective (e.g., benzoyl peroxide, triamcinolone), antibiotics/antimicrobials (e.g., clindamycin, doxycycline, fluconazole, minocycline, rifampin, trimethoprim); 3) vitamin A (tretinoin); 4) medication for mental disorders (e.g., bupropion, paroxetine, and ziprasidone) and smoking cessation (nicotine, varenicline); and 5) other treatments for related comorbidities such as cardiovascular conditions (atorvastatin and heparin), diabetes (dulaglutide, metformin), and pain (capsaicin) (**Fig. 2c**). These associations remained reproducible in the validation analyses (**Methods**, **Extended Data Fig. 2-3** and **Supplementary Tables 7-9**).

### Discovery of phenotype-informed gene module for Hidradenitis Suppurativa (PiHSM)

The PiMInfer framework next integrated the EHR-derived HS comorbidity profiles with a biomedical knowledge graph (BKG) to derive a phenotype-informed HS gene module (PiHSM). Here, we used iBKH^5^, a comprehensive BKG that we previously constructed. First, we mapped the 448 HS-associated comorbidities to disease and symptom nodes in iBKH, defining these as HS-iBKH entry nodes. Second, we assigned each entry node an initial HS risk score, defined as its estimated coefficient in the above association analysis, and then propagated the signals through iBKH’s graph architecture using a random walk with restart^24^, thereby transmitting the phenotypic signals from phenotypes (entry nodes) to other domains in iBKH including the gene nodes. Third, we restricted to gene nodes and assembled the HS gene module using a network approach^25^ that jointly enhances (i) the HS risk over genes at module level and (ii) intra-module connectivity. More details of these procedures can be found in **Methods** and **Fig. 3a**. This yielded a coherent HS gene module, PiHSM (**Fig. 3b**), comprising 69 genes interconnected by 330 interactions. Notably, PiHSM encompasses tumor necrosis factor (*TNF)*, interleukin-1 beta (*IL1B*), interleukin-6 (*IL6*), and interferon gamma (*IFNG*); and they are located at the central of PiHSM with high degree centrality, measuring their connection density to other module genes. This is supported by their well-established roles as key pro-inflammatory mediators in driving the dysregulated immune response in HS lesions^26^, demonstrating the capacity of our approach to detect disease pathological gene signatures.

**Figure 3.**
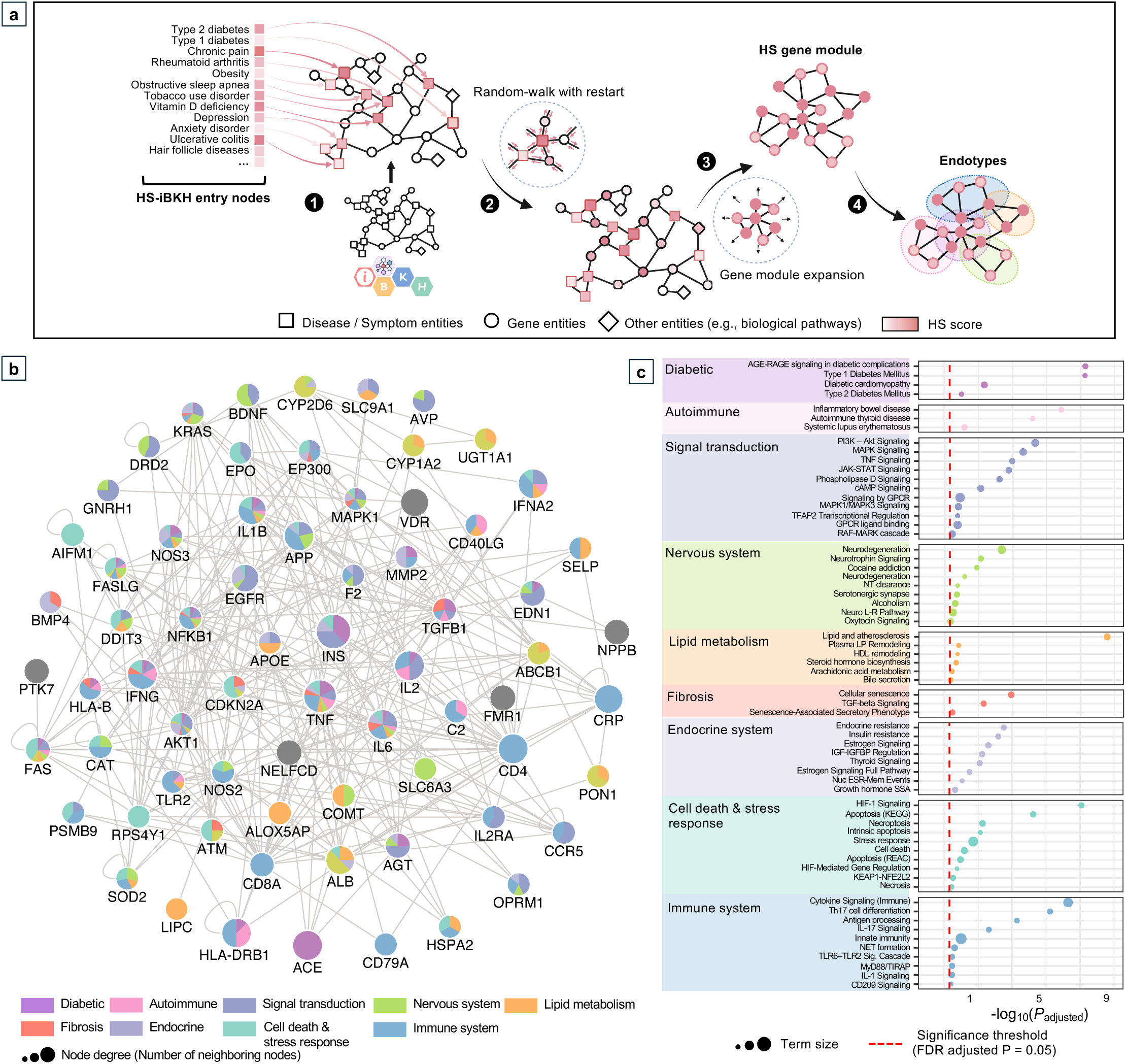
Network analysis integrating phenotypic comorbidity profile with biomedical knowledge graph (BKG) identifies the phenotype-informed molecular network for HS (PiHSM) and functional endotypes. (**a**) An illustration of the BKG-based network approach. Comorbidities significantly enriched in HS from EHR analysis were mapped to corresponding disease and symptom nodes in our BKG, iBKH^5^, defining HS-iBKH entry nodes. Each entry node was assigned an initial HS risk score (association coefficient), indicated as color intensity (step 1). The signals were propagated through the BKG structure using random walk with restart, transmitting phenotypic information to other domains, including genes (step 2). We constructed raw modules using a module-expansion procedure in which genes that increase module-level risk scores and intra-module connectivity were iteratively added. We generated multiple raw modules by repeating the expansion procedure; we then merged the raw modules and retained the largest connected component as the final disease module (step 3). Finally, we subdivided the module into functional endotypes through pathway analysis (step 4). More details can be found in **Methods**. (**b**) The resulting PiHSM. Nodes represent genes, edges denote known biological interactions, node size reflects degree centrality, and pie-chart colors indicate pathway memberships defined in nine pathway categories in (**c**). We further subdivided PiHSM into nine functional endotype subnetworks (**Extended Data Figure 4**).

### PiHSM revealed nine functional endotypes of HS

We next performed pathway enrichment analysis based on the PiHSM genes, and detected the involvement of diabetic, autoimmune, signal transduction, nervous system, lipid metabolism, fibrosis, endocrine system, cell death & stress response, and immune system pathways in HS (**Fig. 3c**). This motivated us to further resolve PiHSM into nine functional endotypes based on genes’ pathway memberships (**Extended Data Fig. 4** and **Supplementary Table 10**), offering finer granularity perspective to elucidate disease mechanisms and improve therapeutic targeting^27,28^. The endotypes are partially overlapping with each other; while the key mediators of HS pathobiology such as *TNF*, *IL1B*, *IL6*, *IFNG*, mitogen-activated protein kinase 1 (*MAPK1*), nuclear factor kappa B subunit 1 (*NFKB1*), and AKT serine/threonine kinase 1 (*AKT1*) participate in multiple functional endotypes. Such a shared, pleiotropic circuitry provides a mechanistic landscape for the marked clinical heterogeneity of HS and its complex comorbidity profile.

### Multiple PiHSM genes were identified to be differentially expressed in HS lesional skin

We sought to validate expression of the PiHSM genes and precisely map the specific cell types and spatial distribution of their potentially altered expression within human HS lesions. To achieve this, we first assembled a comprehensive single-cell RNA sequencing (scRNA-seq) transcriptome profile by integrating our data from the skin samples of HS patients and healthy donors, along with two independent public datasets^11,29^. After data quality control, integration, and processing procedures, we obtained a total of 61,796 cells in HS lesions and 27,989 cells from normal controls, subdivided into eight cell populations including B cell, T cell, keratinocyte, fibroblast, myeloid, plasma, endothelial cell, and myofibroblast (**Fig. 4a-b**, **Extended Data Fig. 5**, and **Supplementary Figs. 1-8**). Through examining their cell-type-specific gene expression profiles, we found that 48 (70%) out of the 69 genes expressed in the HS lesional skins (**Extended Data Fig. 6a**). We next calculated module scores of PiHSM within each cell population in normal and HS lesional skins, respectively, and found that the PiHSM module scores in HS lesions were significantly higher than those in the normal skins (two-tailed t-test estimated *P* values < 0.001) in most cell type (**Fig. 4c** and **Extended Data Fig. 6b**). In addition, cell-type-specific differential expression analyses were performed using the pseudobulk approach^29,30^, to account for dependence between cells from the same individuals. Out of the 48 expressed PiHSM genes, 37 (77%) were differentially expressed (false discovery rate [FDR] adjusted *P* values < 0.05) in HS lesional skins (**Fig. 4d**). Further, Visium spatial transcriptome (10X Genomics) and immunofluorescent (IF) microscopy were performed to confirm their differential expression with spatial information at the RNA and protein level, respectively (**Fig. 4e-h**, and **Extended Data Fig. 6c**). We demonstrated significant upregulations of *TLR2, C2, TGFB1, NFKB1, VDR, APP, SOD2, HLA-B, and PSMB9* in HS lesional skin, and downregulation of *APOE* in HS lesional keratinocytes. Notably, *PSMB9* (proteasome subunit beta type-9) and *HLA-B* showed strongest upregulation in HS lesions in nearly all cell types. Because *PSMB9* plays a critical role in MHC class I antigen presentation machinery, and *HLA-B* is a class I molecule, our findings highlight MHC class I-mediated antigen presentation as a potentially important contributor in HS pathogenesis that has not been previously evaluated and appreciated.

**Figure 4.**
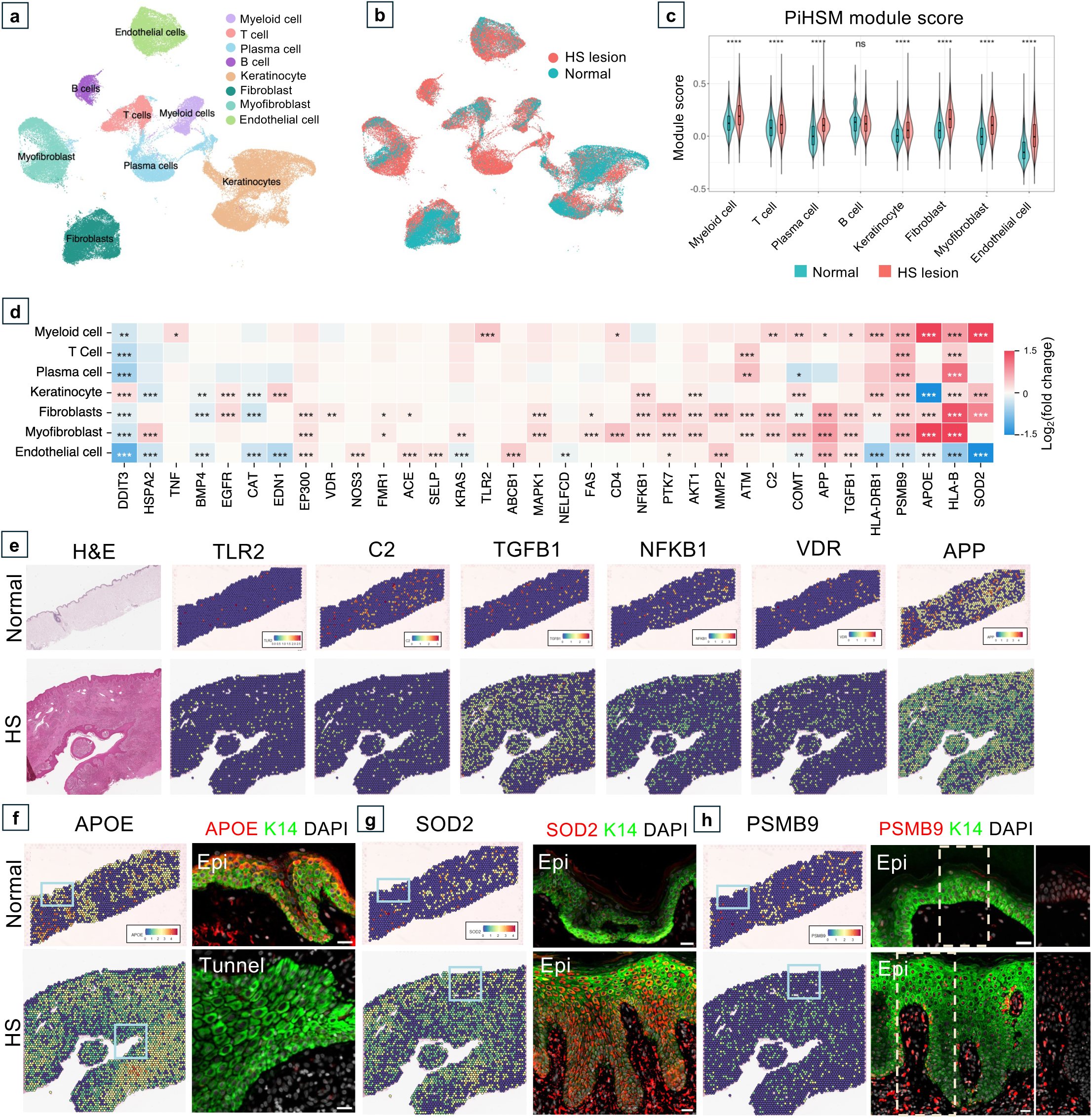
Multimodal biological evidence reveals differential expression of multiple PiHSM genes in HS lesional skin. (**a-b**) Uniform Manifold Approximation and Projection (UMAP) visualization of cell populations from single-cell RNA-seq of HS lesional and normal skin. We integrated the NYU dataset and two publicly available datasets containing normal skin samples (GSE173706 and GSE195452). Cell types were annotated using known marker genes. The UMAP plots are colored by cell types (**a**) and conditions, i.e., HS lesion vs. normal (**b**). (**c**) Comparisons of PiHSM module scores between HS lesions and normal controls across major cell populations. For each cell type, module scores were calculated based on the PiHSM genes that were expressed in ≥ 2% of cells and upregulated in HS lesional skins. (**d**) Heatmap of differentially expressed genes (HS lesion vs. normal) across cell types. Genes upregulated in HS are shown in red, and downregulated genes are shown in blue. *FDR < 0.05; **FDR < 0.01; ***FDR < 0.001. (**e-h**) We integrated two 10x Visium spatial transcriptomics including one normal axilla and one HS patient sample containing tunnel. Spatial feature plots for multiple genes are shown with the same expression scale. Corresponding immunofluorescence staining was performed to evaluate if the protein expression pattern matches its transcription (**f-h**). Designated proteins are visualized as red, with references of keratinocytes by K14 (green) and nuclei with DAPI staining (grey). Scale bars: 25 µm.

### *In silico* drug repurposing based on PiHSM prioritized Carfilzomib as a potential candidate for HS treatment

We further extended PiHSM toward therapeutic discovery for HS (**Fig. 5a**). First, the upregulation of PiHSM module gene *PSMB9* in HS lesional skins prompted us to examine whether suppressing MHC class I antigen presentation may lead to favorable outcomes in HS. To this end, we identified five known *PSMB9* inhibitors, Carfilzomib, Bortezomib, Ixazomib, Marizomib, and Oprozomib, using the Drug–Gene Interaction Database (DGIdb) V5.0^31^, and applied a network-proximity based approach to predict drug-disease associations^32^. Specifically, the interactions between each drug candidate’s target network and PiHSM were assessed, along with its functional endotypes (**Fig. 5a**). For each drug, a set of known physical targets were compiled and calculated for a *z*-score of its network proximity to PiHSM based on 1,000 permutations. A network proximity *z* ≤ - 1.5 indicates a strong association between the drug’s target profile and the disease molecular networks^33^. More details can be found in **Methods**. The capability of the efficacy prediction model was evaluated by testing the two FDA-approved treatments for HS—Adalimumab (TNF-α blockades) and Secukinumab (IL17 blockade). Both drugs showed significant proximity to the PiHSM network (Adalimumab, *z* = -4.33; Secukinumab, *z* = -2.35) (**Fig. 5b**), supporting the potential of using PiHSM-anchored network proximity approach to identify effective therapies for HS.

**Figure 5.**
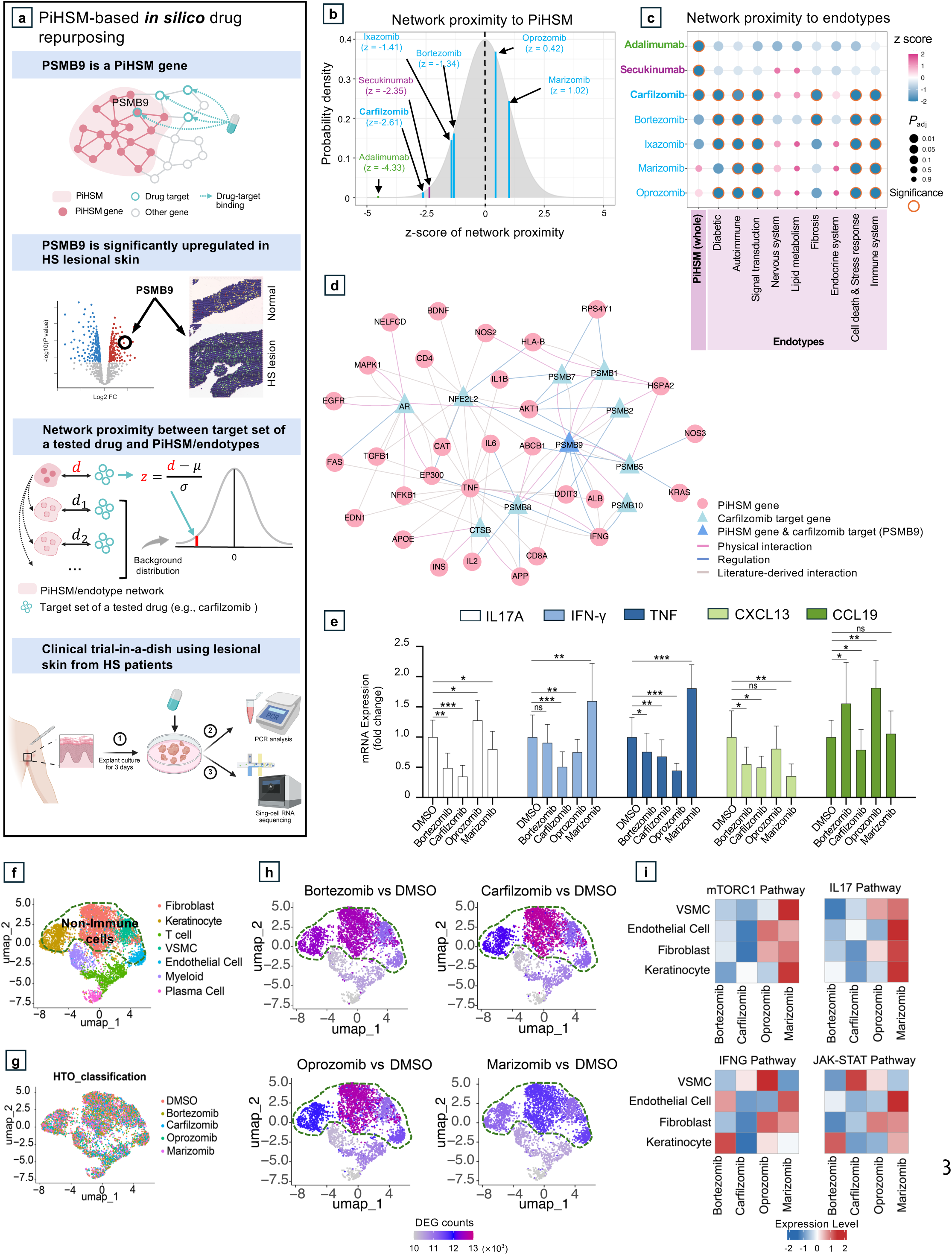
PiHSM-based *in silico* drug repurposing and experimental validation predict Carfilzomib as a promising repurposing candidate for HS treatment. (**a**) Overview of PiHSM-based drug repurposing. First, the marked upregulation of PiHSM module gene *PSMB9* in HS lesions prompted us to investigate if suppressing MHC class I antigen presentation could be therapeutically beneficial for HS. We thus examined five PSMB9 inhibitors including Carfilzomib, Bortezomib, Ixazomib, Marizomib, and Oprozomib. We next applied a network-proximity based approach to assess potential of each drug candidate. Last, we experimentally examined the efficacy of the drug candidates using freshly excised lesional skin from HS patients. Quantitative PCR (qPCR) and single-cell RNA-sequencing analyses were performed to test anti-inflammatory activity within the HS lesion microenvironment. More details can be found in **Methods**. (**b-c**) Network proximity (*z*-scores) of the drug candidates with PiHSM and functional endotype networks. The network proximity (*z*-scores < -1.5) suggest Carfilzomib as a promising repurposing candidate. (**d**) Network visualization illustrating connections between Carfilzomib targets (triangles) within the PiHSM genes (circles). Of note, *PSMB9* is both PiHSM gene and Carfilzomib target. (**e**) qPCR analysis demonstrates that Carfilzomib most effectively suppresses inflammatory marker expression compared to the other four *PSMB9* inhibitors in HS. (**f**) Uniform Manifold Approximation and Projection (UMAP) visualization of PSMB9 inhibitors treated single cell RNA-seq data, color-coded by cell type. The circle-highlighted areas indicate non-immune cells. (**g**) UMAP visualization colored by 5 different hashtags (HTOs) showing demultiplexing assignments. (**h**) UMAP visualizations that illustrate distribution of differentially expressed gene (DEG) counts across cell type clusters under different PSMB9 inhibitor treatments, colored according to DEG counts. The circle-highlighted areas indicate non-immune cells. (**i**) Heatmaps showing the change in Hallmark TNF, IFN-γ, IL-17, mTORC1, and JAK–STAT response module scores in non-T cell populations for each drug relative to control (Treatment − Control). Values are cell-type means, row-scaled (z-score per cell type); blue indicates lower pathway activity while red indicates higher activity under drug. Cell-type labels are on the left. Abbreviation: HS, Hidradenitis Suppurativa.

We next evaluated each PSMB9 inhibitor individually in the network-proximity prediction model. Although all five drugs were known to target *PSMB9*, they showed distinct proximity profiles to the PiHSM network and endotype subnetworks (**Fig. 5b-c** and **Extended Data Fig. 7**). Specifically, amongst all the drugs tested, Carfilzomib exhibited the strongest proximity to the PiHSM network (*z* = -2.61) and most endotype subnetworks including diabetic, autoimmune, signal transduction, fibrosis, cell death and stress response, and immune system (**Fig. 5b-c** and **Extended Data Fig. 7**). Given that Carfilzomib had been previously FDA-approved for multiple myeloma^34^, our results highlight its potential as a repurposing candidate for HS. We further visualized the Carfilzomib targets within the PiHSM neighborhood (**Fig. 5d**). Beyond directly interacting with the PiHSM gene *PSMB9*, Carfilzomib targets including multiple proteasome β- subunits (PSMB family), androgen receptor (*AR*), *NFE2L2* (nuclear factor erythroid 2-like 2), and cathepsin B (*CTSB*) that show extensive interactions with other PiHSM genes, reinforcing its mechanistic relevance to HS pathology.

### Carfilzomib exhibits significant suppression of key inflammatory cytokines in human HS lesional skin

To experimentally examine the efficacy of PSMB9 inhibitors in HS, we obtained freshly excised lesional skin from HS patients, cultured the tissues with each inhibitor for three days, and subsequently analyzed gene expression by quantitative PCR (**Fig. 5a**). We found that Carfilzomib markedly suppressed the expression of key pathogenic cytokines—*IL17A*, *IFN-γ*, *TNF*, *CXCL13*, and *CCL19*, compared to DMSO control and the other PSMB9 inhibitor-treated HS tissues (**Fig. 5e**).

To further delineate the molecular and cellular effects of PSMB9 inhibitors in HS, we performed single-cell RNA sequencing to investigate cell type-specific drug responses in HS lesional skin explants (**Fig. 5a**). Uniform Manifold Approximation and Projection (UMAPs) with annotated clusters and even distribution of cells from each treatment are shown in **Fig. 5f-g**. The number of differentially expressed genes (DEGs) in specific cell clusters after treatment of PSMB9 inhibitors are individually shown in **Fig. 5h**. We found that the most substantial transcriptional changes occurred in non-immune cells, particularly fibroblasts and keratinocytes, whereas immune cells showed much fewer changes (**Fig. 5h**). Pathway module analysis based on DEGs revealed that bortezomib-and carfilzomib-treated HS tissues significantly downregulated mTORC1 and IL17 pathways relative to DMSO control HS tissues. Carfilzomib further demonstrated the most pronounced inhibition of IFN-γ and JAK–STAT signaling in non-immune cell populations (**Fig. 5i**). Together, these findings highlighted Carfilzomib’s superior ability to suppress the inflammatory programs central to HS pathogenesis within the lesional tissue microenvironment.

### Potential combination therapy of Carfilzomib with approved HS medications

Beyond single-agent proximity analyses, we next evaluated the potential of combining Carfilzomib with the FDA-approved HS treatments Secukinumab and Adalimumab. Network visualizations revealed that Carfilzomib shares no direct targets with either drug; and their target footprints show minimal interaction within the interactome (**Fig. 6a-b**). To quantify functional overlap between two drugs, we calculated a network separation score based on the topological distance between target modules of the drugs within the human interactome (**Methods**)^35^. A separation score > 0 indicates that two agents occupy distinct network neighborhoods, reflecting pharmacological complementarity and potential for synergistic benefit^35^. As shown in **Fig. 6c**, Carfilzomib demonstrates functional complementarity with both Secukinumab (separation score = 1.52) and Adalimumab (separation score = 0.52). Together, these results suggest that combining Carfilzomib with existing HS therapies may enhance therapeutic efficacy, though experimental validation will be essential to confirm these predictions.

**Figure 6.**
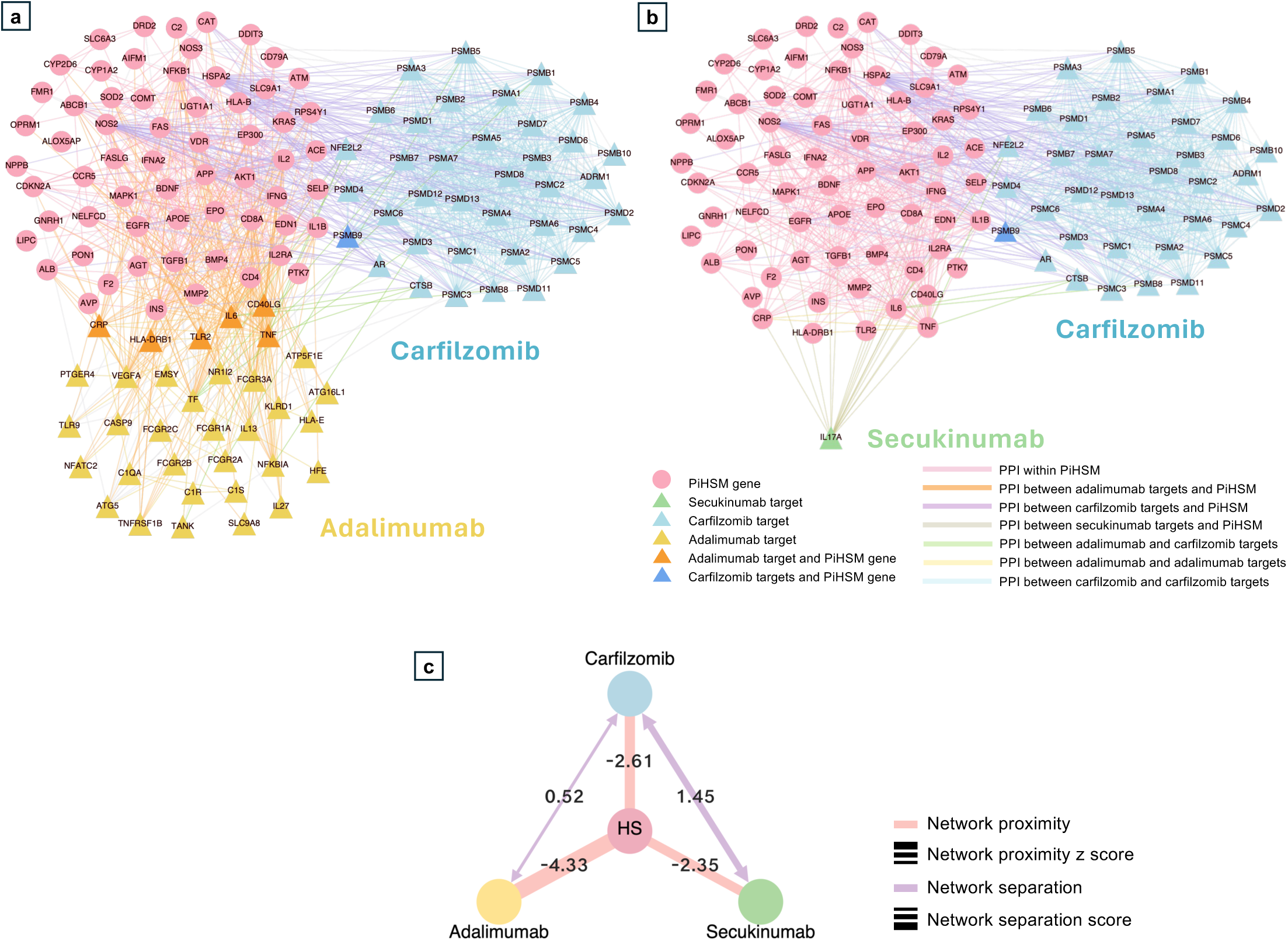
PiHSM-based drug combination analysis for Hidradenitis Suppurativa (HS). (**a-b**) Network visualizations showing relationships between PiHSM and the target modules of Carfilzomib and approved HS medications Adalimumab (**a**) and Secukinumab (**b**), respectively. (**c**) Conceptual diagram summarizing relationships between PiHSM and target modules of Carfilzomib and then approved HS medications Adalimumab and Secukinumab. A network proximity z-score < –1.5 indicates potential therapeutic relevance of a drug for HS based on PiHSM, whereas a separation score > 0 between two drugs reflects pharmacological complementarity and potential synergistic benefit.

## Discussion

Computational drug repurposing has produced notable successes and is rapidly emerging as a prominent direction in modern therapeutic discovery, rising alongside recent advances in artificial intelligence (AI) and machine learning (ML)^36–41^. However, most of the prior efforts have been constrained by incomplete biological data, the lack of suitable disease models for complex diseases, and low translational success due to insufficient experimental or clinical validation^37,42^. Clinical heterogeneity further compounds these challenges and is widely recognized as a major barrier to effective drug discovery^37,43,44^. To address these, our PiMInfer framework leverages the cross-domain relational nature of biomedical knowledge graphs (BKGs) to translate RWD-derived deep phenome-wide comorbid architecture of a human disease into its molecular mechanisms, thereby accelerating therapeutic identification. As global efforts continue to accumulate and integrate increasingly large-population real-world patient data^45^, coupled with computational power and AI/ML methodological advances, the scale and diversity of patient records accessible to PiMInfer will continue to expand, significantly surpassing that of genetic and multi-omic studies, facilitating rapid and more effective drug repurposing.

While EHRs provide rich and temporally resolved phenotypes enabling phenome-wide deep phenotyping^45^, existing EHR-based approaches typically remain restricted to comorbidity associations^46–48^, lacking depth at the molecular mechanisms such as disease pathogenesis and target biology. Our PiMInfer largely strengthens the power of EHRs in drug development through introducing a unified, cross-modal inference pipeline that systematically projects EHR-derived disease comorbidity profiles onto the topological structure of a high-quality BKG, enabling seamless information flow from clinical phenotypes and molecular mechanisms. Methodologically, PiMInfer integrates a BKG-based signal propagation with a network module identification approach that jointly optimizes clinical coherence and molecular connectivity. The resulting disease modules thus exhibit markedly improved mechanistic interpretability and stability, providing a principled foundation for therapeutic target discovery and drug repurposing. Compared with omics-driven approaches (e.g., GWAS, scRNA-seq), PiMInfer delivers a substantial and complementary methodological advancement by enabling multi-scale, clinically anchored inference of actionable molecular signatures and functional endotypes – particularly valuable for rare or understudied population of complex diseases that lack large-scale, high-quality omics data.

We applied our PiMInfer framework to HS, as an example of a debilitating disease with complex comorbidities and limited effective treatments. In contrast to better-studied comorbid conditions such as rheumatoid arthritis, inflammatory bowel disease, and diabetes, far less biological and clinical information is available for HS. Leveraging large-scale EHR data and biological insights from these better-characterized comorbidities, we developed PiHSM to enable deeper investigation of molecular mechanisms underlying HS pathogenesis. Notably, TNF occupies a central hub in the PiHSM network, with the highest connectivity to other module genes, underscoring its central role in HS and aligning with the moderate efficacy observed for anti-TNF therapy, the first FDA-approved treatment for HS^49^. Remarkably, nearly all targets of drugs currently in HS clinical trials are represented in PiHSM, including *IL1B* (Lutikizumab)^50^; *NFKB1* (downstream of IL-36R targeted by Spesolimab)^51^; *C2* (in the complement pathway, alongside C5A targeted by Vilobelimab)^52^; *IL2*, *ILRA*, *IL6*, *IFNA2*, *IFNG*, and *EP3* (associated with the JAK/STAT pathway for Povorcitinib^53^ and Upadacitinib^54^); *TLR2* and *IL1B* (upstream of IRAK4 for SAR444656)^55^; *IL6* (downstream of IL17A/F for Bimekizumab)^56^; and *CD40LG* and *CD79A* (upstream of BTK for Remibrutinib)^11,57^, providing supporting evidences that genes included in PiHSM network may play critical roles in HS pathogenesis and possess the potential to serve as drug targets for HS.

We prioritized *PSMB9* from PiHSM because it is significantly upregulated across nearly all cell types in lesional skin in our multimodal validation and, importantly, was also implicated by a recent GWAS study of 6,361 HS cases^58^. Among the five PSMB9 inhibitors tested, Carfilzomib demonstrated the most robust immune suppression, consistent with PiHSM-based drug response prediction. Our scRNA-seq analyses of Carfilzomib-treated HS tissues further revealed marked downregulation of IL17, mTOR, and IFN-γ signaling pathways in non-immune cells within the HS lesional microenvironment, underscoring the pathogenic roles of IFN-γ-induced antigen presentation in HS and suggesting its potential as a combinatorial candidate to enhance the efficacy of currently approved therapies.

Limitations of the study include the following. First, although routine-care EHRs provide rich longitudinal phenotypic information, issues including data sparsity, missingness, coding inconsistency, and site-specific practice patterns may affect disease phenotyping and comorbidity profiling. Even though the EHR repositories used here adopt the Observational Medical Outcomes Partnership (OMOP) Common Data Model (CDM), institutions without a standardized CDM face additional challenges in mapping phenotypes (e.g., diagnosis codes) to BKG nodes, increasing the risk of alignment errors and information loss. Second, despite our BKG (iBKH) integrated data from diverse curated sources^5^, biomedical knowledge may be incomplete and unevenly annotated, necessitating ongoing updates to maintain coverage and accuracy. Third, as this study focused on HS as a proof of concept, diagnostic delays and under-recognition of HS may introduce left-censoring and under-ascertainment in EHR-derived cohorts. Finally, our drug repurposing predictions are *in silico*–driven and supported by ex vivo assays using human lesional skin. To study other diseases where patient tissues are not readily available, additional validation using animal models or organoid systems may be required, though each has limitations^59^.

In conclusion, we present a generalizable framework capable of disentangling heterogeneous comorbidity profiles to delineate disease gene modules and prioritize drug repurposing candidates for human complex diseases. Our findings and experimental validation using HS as an example of understudied complex diseases demonstrated the strength and accuracy of PiHSM in therapeutic prioritization for HS and emphasized crucial insights: monotherapy alone may be insufficient to manage a complex, multifactorial disease like HS, and a strategic approach involving rational combination treatments will be essential to achieve more durable and favorable patient outcomes.

## Data Availability

Data from the All of Us Research Program were accessed through the All of Us Researcher Workbench (https://workbench.researchallofus.org/) under the standard application procedure. Data from the INSIGHT Clinical Research Network were obtained under institutional agreement. INSIGHT data can be requested at https://insightcrn.org/ upon approval. Single-cell and spatial transcriptomics data generated by our team are publicly available at Gene Expression Omnibus (GEO) under accession number GSE158955 (data from 9 samples are currently available, with the remaining samples to be released upon acceptance). Publicly available single-cell RNA sequencing datasets were obtained from the GEO under accession numbers GSE173706 and GSE195452 (https://www.ncbi.nlm.nih.gov/geo/). Drug–target interaction data were retrieved from DGIdb V5.0 (https://dgidb.org)^31^, DrugBank (https://go.drugbank.com/), BindingDB (http://www.bindingdb.org/), ChEMBL (https://www.ebi.ac.uk/chembl/), the Therapeutic Target Database (https://idrblab.net/ttd/), PharmGKB (http://www.pharmgkb.org/), and the IUPHAR/BPS Guide to PHARMACOLOGY (https://www.guidetopharmacology.org/). The BKG (iBKH) used in this study is available at https://github.com/wcm-wanglab/iBKH.

## Code Availability

All analyses were conducted in Python 3.7 and R version 4.1. Logistic regression models were built using ‘glm’ function in base R. Propensity score matching was performed using the MatchIt 4.7.2 R package. All other statistical tests were performed in R. Single-cell and spatial transcriptomics analyses were performed using the Seurat V5 package^60^. Network visualization was done using Cytoscape V3.10.2 (https://cytoscape.org/index.html)^61^. Other data visualization, including bar plots and forest plots were generated using the ‘matplotlib 3.0’ package in Python (https://matplotlib.org).

All computer code for this analysis is available at https://github.com/changsu10/HS_phenotyping_drug_repurposing

## Data Availability

The data used in this study are available through controlled-access platforms including the All of Us Research Program Researcher Workbench and INSIGHT Clinical Research Network for eligible researchers.

https://workbench.researchallofus.org/

https://insightcrn.org/

https://github.com/wcm-wanglab/iBKH

https://www.ncbi.nlm.nih.gov/geo/

## Author Contributions

C.S. and C.P.L conceptualized the study and designed the experiments. C.S., C.P.L., W.-T.W., M.Z, J.T., and F.W., interpreted the data and wrote the manuscript, with input from all co-authors. C.S., C.P.L. and F.W. supervised the research, with C.S. and C.P.L. leading the study. W.-T.W. conducted the primary analysis in EHRs. W.-T.W. and Z.X. conducted validation analysis in EHRs. M.Z. and W.-T.W. conducted biomedical knowledge graph (BKG) analysis. M.Z., A.K., and M.W. conducted single-cell RNA-seq analysis. E.S.C. performed surgeries and patient care. J.T. procured and processed patient samples. M.-J.L. performed spatial data analysis and IF imaging. M.Z. and A.K. performed in silico drug repurposing. J.T. performed human patient lesional skin explant experiments for treatment efficacy testing. H.T., A.P., K.T.H., S.R.C, and L.P. provided critical comments and interpretations.

## Acknowledgement

C.S. is supported by NIH R01 grant 1R01NS140142. C.S. also received startup funds and funds for Walsh McDermott Scholar in Public Health from the Department of Population Health Sciences of Weill Cornell Medicine, Cornell University.

## Methods

### Deep phenotyping – phenotypic association analysis based on EHRs

#### Study cohort

We used patient-level clinical data from two large-scale real-world EHR repositories:

1. The All of Us (AoU) Research Program^62^, a nationwide initiative in the United States that collects diverse, longitudinal health data from over one million participants to advance precision medicine and population health research. We analyzed the data using the AoU Research Workbench, which allows authorized users to conduct research projects without requiring individual Institutional Review Board (IRB) review. We used de-identified, longitudinal EHRs of participants in the AoU database with a cutoff date of July 1, 2022.
2. The INSIGHT Clinical Research Network (CRN)^63^, supported by the Patient-Centered Outcomes Research Institute (PCORI), covering longitudinal clinical records for over 15 million patients in the five top academic medical centers in NYC metropolitan area, including Weill Cornell Medicine (WCM), Albert Einstein School of Medicine/Montefiore Medical Center, Columbia University Irving Medical Center, Icahn School of Medicine/Mount Sinai Health System, and NYU Grossman School of Medicine/NYU Langone Health. We used de-identified, longitudinal EHRs of patients in INSIGHT CRN between general 2010 to September 2024. Use of INSIGHT CRN data was under the approval of IRB at WCM (IRB number 24-08027797).

The study populations were limited to individuals aged 18 to 90. Individuals whose date of birth and sex information are missing and whose EHR records are shorter than 1 year (insufficient records) were excluded for analysis. The HS population were defined using at least one of the International Classification of Diseases, Ninth & Tenth Revision (ICD-9 & ICD-10) codes including 705.83 (ICD-9) and L73.2 (ICD-10), and the Systematized Nomenclature of Medicine (SNOMED) codes including 18638007, 59393003, 69741000, 402826001, and 402828000. A prior study has validated that this algorithm can achieve a positive predictive value of 79.3% and an accuracy of 90% for diagnosis of HS^64^. In AoU, all remaining participants who had no HS diagnosis were used as control individuals. In INSIGHT CRN, we randomly sampled 565,380 patients who had no HS diagnosis or any diagnoses listed in the predefined whitelist. Workflow for constructing the study cohorts were illustrated in **Extended Data Fig. 1**.

#### Clinical variables

The AoU EHR utilizes the SNOMED and ICD diagnosis codes and INSIGHT uses ICD diagnosis codes. Thus, we first normalized the records to ICD-10-CM codes using the SNOMED CT to ICD-10-CM Map downloaded from the Unified Medical Language System (UMLS)^65^. All ICD-9 and ICD-10 codes were mapped to phecodes using the Phecode Map Version 1.2^66^. We also studied medication prescription records of participants. Medications were standardized to generic names of active ingredients using RxNorm^67^.

#### Association analyses

We used multivariate logistic regression models to test the association between HS and each phecode, adjusting for covariates including age, sex, race, and ethnicity. Similarly, we also fitted multivariate logistic regression models to test the association between HS and each medication ingredient, adjusting for covariates as mentioned above. Significant associations were defined based on Bonferroni-corrected threshold (i.e., *P* value < 0.05 / number of variables). We reported the estimated coefficients (𝛽) to quantify the strength of the associations.

#### Validation analyses

To assess the robustness of our identified phenotypic associations, we replicated the analysis by comparing HS patients with 1:10 demographic (age, sex, race, and ethnicity)–matched controls in AoU. Matching was conducted using a propensity score matching (PSM) approach, implemented via the MatchIt R package^68^. We further validated our findings in the INSIGHT data by comparing HS patients with both all available controls and a 1:10 demographic-matched control group constructed using the same matching procedure.

### Biomedical knowledge graph (BKG)–based approach for disease molecular network identification

#### iBKH

We have constructed a comprehensive BKG, termed the integrative biomedical knowledge hub (iBKH)^5^. As of now, iBKH has integrated and harmonized data from 18 publicly available biomedical knowledge sources, including biomedical ontologies, manually curated knowledge bases, existing biological knowledge graphs, and other biomedical data. It includes approximate 2.4 million biomedical entities (nodes) of 11 types, such as genes, diseases, symptoms, pathways, drugs, etc.) as well as over 48 million relations of 45 relation types, such as disease-associated-with_gene, disease-associated-with_disease, gene-bind-gene, etc.).

#### Gene module identification

To eliminate noise from irrelevant information^69^, we extracted a subgraph with core biomedical knowledge from iBKH. Specifically, we included key entity types including diseases, symptoms, genes, and biological pathways, as well as relations among them. This resulting BKG consists of 110,992 nodes and 1,054,557 relations, forming the basis for downstream network analysis for gene module discovery.

We next linked the identified HS comorbidities (i.e., HS-associated phecodes that met the Bonferroni-corrected significance threshold, *P* value < 0.05/total number of phecodes) into iBKH. iBKH contains ICD codes and UMLS Concept Unique Identifiers for the disease and symptom nodes, thus we mapped these phecodes to iBKH nodes using Phecode Map Version 1.2^33^. To enhance mapping accuracy and coverage, we also utilized the unified medical language system (UMLS)^65^. The resulting mapped disease and symptom nodes were designated as HS-iBKH entry nodes. Note that one phecode can correspond to multiple HS-iBKH entry nodes.

We employed a network medicine approach^25,70,71^ to derive the gene module for HS using the comorbidity signals. The approach demands two inputs, the BKG (i.e. iBKH) we constructed and the HS-iBKH entry nodes along with their corresponding association effect 𝛽. First, we initialized a node-level disease (HS) risk score (𝑖) for each node 𝑖 in the BKG: for each HS-iBKH entry node, 𝑠(𝑖) = |𝛽|; for all other nodes, 𝑠(𝑖) = 0. Next, we propagated these risk scores through the BKG network using a random walk procedure with restart to obtain a smoothed risk score *ŝ*(𝑖) for each node 𝑖. Such a procedure enables propagating the EHR-derived signals (HS risk scores) from the phenome to genome within the BKG. Next, we retained all gene nodes (along with their scores *ŝ*(𝑖)) and relations among them. We utilized an iterative expansion process below to construct a module 𝑀. Specifically, a module 𝑀 was initialized with a randomly selected HS entry node. At each step, a neighboring gene node 𝑗 ∈ 𝛤*_M_* (i.e., connected to any node in 𝑀) was considered for inclusion if it satisfied two criteria: (a) *P_connectivity_*(𝑀 ∪ {𝑗}) < 0.05 (calculated with **Eq. 1**), indicating that the resulting module remains significantly more densely connected internally than expected by chance; and (b) 𝑆(𝑀 ∪ {𝑗}) > 𝑆(𝑀) (calculated with **Eq. 2**), indicating that module-level risk score increased upon involving gene 𝑗). That said, the module was constructed by simultaneously enhancing network cohesion (connectivity) and aggregating disease relevance (HS risk score).

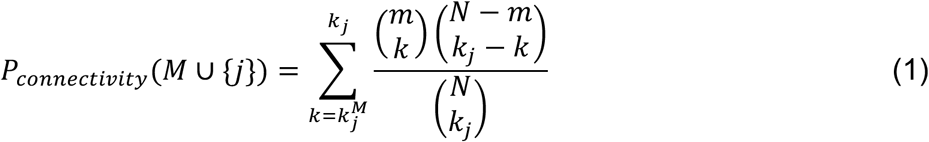

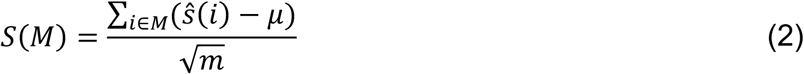

Here, 𝑚 denotes of current number of nodes within 𝑀, 𝑁 is the total number of nodes within the BKG network, 𝑘_*j*_ is the degree of node 𝑗 (i.e., number of neighbors of node 𝑗 within the graph), 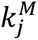 is the number of nodes within 𝑀 that link to, *j*, 𝜇 denotes the averaged risk score of all nodes within 𝑀. We terminated module expansion when *P_connectivity_*(𝑀 ∪ {𝑗}) ≥ 0.05 or 𝑆(𝑀 ∪ {𝑗}) cannot be increased when involving any new node 𝑗. We repeated the above procedure 100,000 times and obtained a collection of raw modules. These modules were ranked in descending order by their module scores, and the top 100 modules (top 0.1%) were aggregated. We obtained the largest connected component to define the final putative HS gene module.

#### Pathway enrichment analysis and endotype classification

We performed pathway enrichment analysis with g:Profiler^72^ using the Kyoto Encyclopedia of Genes and Genomes (KEGG)^73^ and Reactome^74^ pathway databases. Pathways with an FDR-adjusted *P*<0.05 were considered significant. Next, we grouped these enriched pathways into functional categories. The gene module was partitioned into multiple functionally coherent subnetworks, each representing a distinct disease endotype. Notably, genes involved in multiple pathways may contribute to more than one endotype, reflecting the pleiotropic nature of gene function.

### 10x Single-cell RNA sequencing

Patient samples were collected from HS cases screened by the Hansjorg Wyss Department of Plastic Surgery at NYU. Protocols for sample collection were reviewed by the IRB at NYU Langone (IRB number: s19-00841). Lesional and perilesional skin, characterized by nodules, abscesses, and draining fistula, were surgically excised from axillary and groin regions. Samples were collected from 14 HS patients, including lesional and perilesional areas of the groin and axilla. Among these patients, 3 had two samples collected at different visits, while the remaining had a single sample. Additionally, two samples were collected from individuals without HS. Entire excisions were collected from human patients and placed directly into sterile Dulbecco’s Phosphate Buffered Saline (DPBS, 1X w/o Ca2+ Mg+) on ice within an hour of the surgery. Areas of interest, focusing on sinus tracts and inflamed epidermis, as well as perilesional areas, were dissected out of the larger sample to 1cm biopsies. These biopsies were then dissociated into a single cell suspension using an enzyme mix of Hanks’ Balanced Salt Solution (HBSS, 1X w/o Ca2+ Mg+), Collagenase at 2 mg/ml, and Liberase TL at 20 mg/ml. The samples were incubated at 37°C on a shaking plate at 100 rpm. After 1 hour, the samples were agitated with a scalpel to further the digestion process. The samples were incubated for 30 more minutes, of which the final 10 minutes were agitated again using a scalpel and Trypsin/EDTA was added to finalize the digestion process. The suspension was then washed in 4% FBS/DPBS and filtered through a 40 mm cell strainer and spun down at 300 g at 4°C for 10 minutes. Red blood cell lysis using ACK Lysis Buffer was then performed, then cells were washed and spun as mentioned before obtaining a cell count using a hemocytometer. PBMC were isolated from blood samples using ACK Lysis Buffer protocol.

Single cell suspensions of approximately 1.0e6/ml in PBS FBS were submitted to the Genome Technology core facility at NYU Langone. The following supplies were used at the manufacturer’s instructions for scRNA-seq: Chromium Single cell 3’ Library and Gel Bead Kit v2 Chromium Single Cell 3’ Library & Gel Bead Kit v2 (PN-120237), Chromium Single Cell 3’ Chip kit v2 (PN-120236_ and Chromium i7 Multiplex Kit (PN-120262). Once the cDNA libraries were generated, they were subject to HiSeq 2500 (illumina) sequencing. Cell libraries were run for 5’ gene expression data.

More details of our scRNA sequencing protocol can be found elsewhere^13^.

### External scRNA-seq data

We also utilized two scRNA-seq resources publicly available from the NCBI Gene Expression Omnibus (GEO; https://www.ncbi.nlm.nih.gov/geo/) database^75^:

#### GSE173706^11^

This study aimed to investigate cell type composition and transcription changes in psoriasis skins comparing to normal skin from healthy donors. scRNA-seq data of normal skin tissues from the healthy donors from this dataset (N = 8) were included for analysis.

#### GSE195452^29^

This study recruited patients who were diagnosed with systemic sclerosis (SSc) and healthy donors to establish a high-resolution atlas of the sclerodermatous skin spectrum for advancing biomarker and therapeutic development in SSc. We used scRNA-seq data of normal skin tissues from 55 healthy donors from this dataset.

### scRNA-seq data analysis

scRNA-seq data analysis was performed using the Seurat V5 package^60^. Cells with fewer than 500 detected genes or more than 20% mitochondrial gene expression were excluded to ensure adequate representation of both genes and cells. Data were normalized using the NormalizeData function, which divides feature counts by the total counts for each cell, followed by a natural logarithmic transformation. Variable features were identified using the FindVariableFeatures function, and gene-level scaling was performed with the ScaleData function. Principal component analysis (PCA) was conducted using the RunPCA function. To integrate data cross samples and datasets while accounting for batch effects, we utilized the IntegrateLayers function with the “Harmony” method^76^ based on the first 50 principal components. The first 30 Harmony embeddings of cells were used to find cell neighbors and clusters using the FindNeighbors and FindClusters (resolution = 0.8) functions, respectively. Cluster-specific markers were identified through using the FindMarkers function. We further used the UMAP^77^ to reduce data dimensionality with the RunUMAP function, for data visualization purposes. We conducted differential expression analysis (DEA) using the psudobulk approach, accounting for the dependence between cells from the same sample^30^, using the AggregateExpression function in Seurat. We performed cell type-specific DEA using DESeq2 comparing HS and normal conditions^78^, accounting for batch effects across samples. An FDR adjusted *P* value < 0.05 was considered significant. We calculated the module score using the AddModuleScore function in Seurat.

### Spatial transcriptomics analysis

Formalin fixed paraffin embedded specimens from HS lesional samples were sectioned onto Visium Spatial Gene Expression Slides (10X Genomics) by the Experimental Pathology Core at NYU Langone (IRB number: s19-00841). Sequencing was performed by the Genome Technology Core at NYU Langone. Space-ranger output files, including the H5 file and relevant sequencing outputs, were used to compile objects in R software according to the Seurat spatial workflow. Data was normalized using SC-Transform before UMAP clustering and PCA analysis. Pathological analysis was used to determine the lymphoid aggregate region on H&E sections of the sample, which was manually selected for using the 10X Genomics Loupe browser v.4.0. The barcodes of the aggregate areas and non-aggregate areas were extracted as csv files and used for comparison by the SetIdent function in Seurat.

### Immunofluorescent (IF) staining and imaging

Patient skin samples from surgical excision were dissected to fit into Tissue-Tek cryomolds and frozen in Tissue-Tek OCT on dry ice. OCT blocks were sectioned at 10 mm onto microscopic slides. Skin sections were fixed using cold acetone for 10 min at 4C. Staining protocol was as previously described^79^. Primary antibodies, as well as pre-conjugated patient-derived antibodies, were incubated with the tissue sections overnight at 4C. Cytokeratin 14 (Biolegend, 1:800), Loricrin (Abcam, 1:50). Secondary antibodies antichicken IgY (1:500) and anti-rabbit IgG (1:800) were incubated for one hour at room temperature. For direct IF, fluorophore-conjugated anti-human IgG1 antibody (Thermofisher, A-11014, 1:500) was used to detect endogenous IgG1. For indirect IF, HS patient sera were used at a dilution of 1:5 and incubated overnight; after washing with PBS 3 times, fluorophore-conjugated anti-human total IgG antibody (Thermofisher, A-11014, 1:500) were incubated for one hour at room temperature. Imaging was performed using the Zeiss 880 confocal microscope at 20x and 63x objective lenses and processed using Fiji (ImageJ).

### *In-silico* drug repurposing

#### Drug-target network

We used a comprehensive physical drug-target interaction network for FDA-approved drugs reported in previous studies^33,80,81^. Specifically, the drug-target interactions were collected and integrated from diverse data sources including DrugBank^82^, BindingDB^83^, ChEMBL^84^, Therapeutic Target Database^85^, PharmGKB^86^ and IUPHAR/BPS Guide to PHARMACOLOGY^87^. A physical drug-target interaction was defined based on binding affinities Ki (inhibition constant/potency), Kd (dissociation constant), IC50 (median inhibitory concentration), or EC50 (median effective concentration) ≤ 10 μM.

#### Network proximity calculation for drug repurposing

Following previous studies^33,80,81^, we defined the average shortest path distance 𝑑(𝑆, 𝑇) between a disease module 𝑆 (i.e., HS molecular module) and the set of targets 𝑇 of a specific drug, as follows:

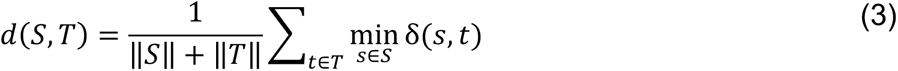

where, ‖𝑆‖ and ‖𝑇‖ denote size of 𝑆 and 𝑇, respectively; 𝛿(𝑎, 𝑏) indicates the shortest path between gene/protein 𝑎 and target 𝑏, within the protein interactome network extracted from iBKH. A permutation test was used to evaluate significance of network-based distance between the HS molecular module we found and each tested drug candidate. Specifically, we first generated a reference distance distribution corresponding to the expected distance between the tested drug’s target set 𝑇 and a random drawn gene set 𝑆*_r_* with the degree (number of neighbors of a gene in the network) distribution and size of HS molecular module 𝑆. We repeated the procedure 1,000 times to obtain a set of 𝑆*_r_*. A z-score was calculated by using the mean and standard deviation of the reference distribution to normalize the observed distance between 𝑆 and 𝑇.

#### Network-based drug combination calculation

To quantify functional overlap between two drugs, we measured overlap of target modules of the two drugs by employing the separation score^35^:

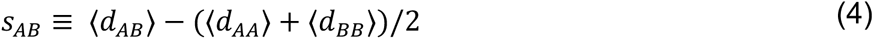

where ⟨𝑑*_AA_*⟩ and ⟨𝑑*_BB_*⟩ are the mean shortest distance among targets within each drug, and ⟨𝑑*_AB_*⟩ is the mean shortest distance between A-B target pairs. According to Cheng et al.^35^, two drugs whose footprints are topologically separated, i.e., 𝑠*_AB_* > 0, are probably pharmacologically distinct and functionally complementary; thus, drug 𝐴 and B are considered as an effective combination if 𝑑(𝑆, 𝐴) < 0, 𝑑(𝑆, 𝐵) < 0, and 𝑠*_AB_* > 0, where 𝑆 denotes the disease molecular module.

### Treatment efficacy testing using human patient lesional skin explants

#### HS explant culture

HS samples were procured in the operation room using a scalpel. Immediately post-procurement, all the samples were soaked into a pre-labelled sterile transport jar which containing transport medium and carefully sealed. The HS sample was dissected into 2 mm wide fragment, and all the fat parts were removed by using sharp sterile blade and forceps. Each fragment was then flattened and placed in a 100 mm tissue culture Petri dish with the epidermal layer facing up. The specimens were allowed to dry for 5 min to immobilize the attachment of all fragments to dish surface before adding medium. The medium used for HS explant culture is mesenchymal stem cell medium (PT-3001, Lonza), which includes both the basal media (PT-3238, Lonza) and the necessary supplements (PT-4105, Lonza) for cell proliferation. Medium was carefully added from the dish edge to barely cover (5-7 ml) the explant’s surface, and then the dish was placed in a 37 °C, 5% incubator overnight. All the treatments were added into the medium and the samples were collected and snap-frozen after 3 days of treatment.

#### Quantitative RT-PCR

Tissue frozen in liquid nitrogen upon sample collection was homogenized using nitrogen-frozen mortar and pestle and resuspended in Trizol. Primary fibroblasts between 1-2 passages were treated with recombinant proteins for an overnight period, then resuspended in Trizol. RNA isolation was performed using Zymo Direct-zol RNA Microprep kit. cDNA was generated using the SuperScript IL VILO system. Melt curve analysis was used to confirm generation of gene targets (**Supplementary Table 11**). mRNA expression levels were normalized by the delta-delta Ct method to housekeeping gene *HPRT1*.

#### DEG counts visualization

We generated a UMAP DEG counts map by forming pseudo-bulk profiles per cell type within each condition: raw RNA counts from Seurat were summed across cells of the same cell type, library sizes were computed as column sums, and values were converted to CPM. Lowly expressed genes were filtered (mean CPM ≥ κ), and per-gene effect size was computed as log2FC (Drug vs Control) with an ε=1 CPM offset for stability. For each cell type, the DEG counts was defined as the number of genes with and mean CPM ≥ κ. This count was assigned to member cells and visualized on the UMAP; for display we optionally clipped values to the 5^th^–95^th^ percentiles to enhance contrast. We then summarized drug effects by cell type by computing per-cell Milo neighborhood effect scores (miloR), averaging within each cell type × condition, and subtracting the matched control mean to obtain a cell-type × drug matrix.

#### Signaling pathway activity analysis

To quantify the activity of key inflammatory signaling pathways (mTOR, IL-17, IFN-γ, JAK/STAT, and TNF) across distinct cell populations, we used log-normalized single-cell RNA-seq expression matrices obtained from Seurat objects. Gene sets corresponding to each pathway were retrieved from the MSigDB Hallmark and Reactome collections via the msigdbr package in R. For each pathway, module scores were calculated at the single-cell level using AddModuleScore function in Seurat, yielding per-cell pathway activity indices.

Mean pathway scores were computed for each cell type and experimental condition, and differential pathway activity was quantified as the difference between drug-treated and control samples. For these signaling, we additionally incorporated ligand–receptor expression analysis by measuring the cytokine and its receptors’ expression to infer sender (cytokine-producing) and recipient (receptor-expressing) cell interactions.

The resulting Δ-pathway scores and receptor expression changes were visualized as clustered heatmaps to depict cell-type-specific responses to treatment and to rank compounds according to their ability to suppress pro-inflammatory signaling.

## Extended Data Figures

**Extended Data Figure. 1.**
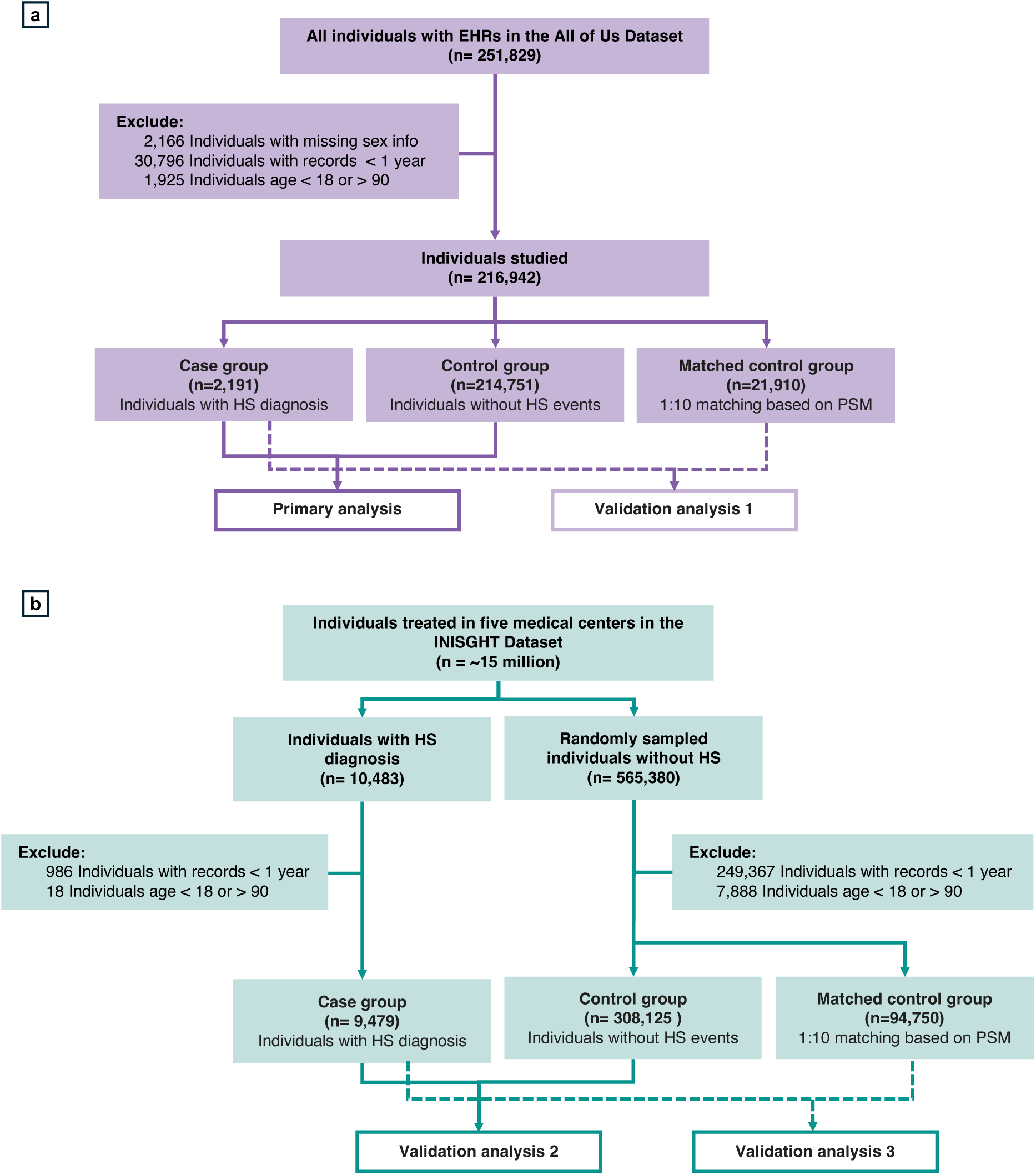
Flowchart of participants included in the study cohort. The flow diagrams illustrate the steps used to construct the analysis cohorts in the All of Us Research Program (**a**) and INSIGHT Clinical Research Network (**b**) according to predefined inclusion and exclusion criteria. In All of Us, participants without electronic health records (EHRs) were excluded. The Hidradenitis Suppurativa (HS) population was defined as individuals with at least one of the following diagnostic codes: ICD-9 705.83, ICD-10 L73.2, and SNOMED codes 18638007, 59393003, 69741000, 402826001, and 402828000.

**Extended Data Figure 2.**
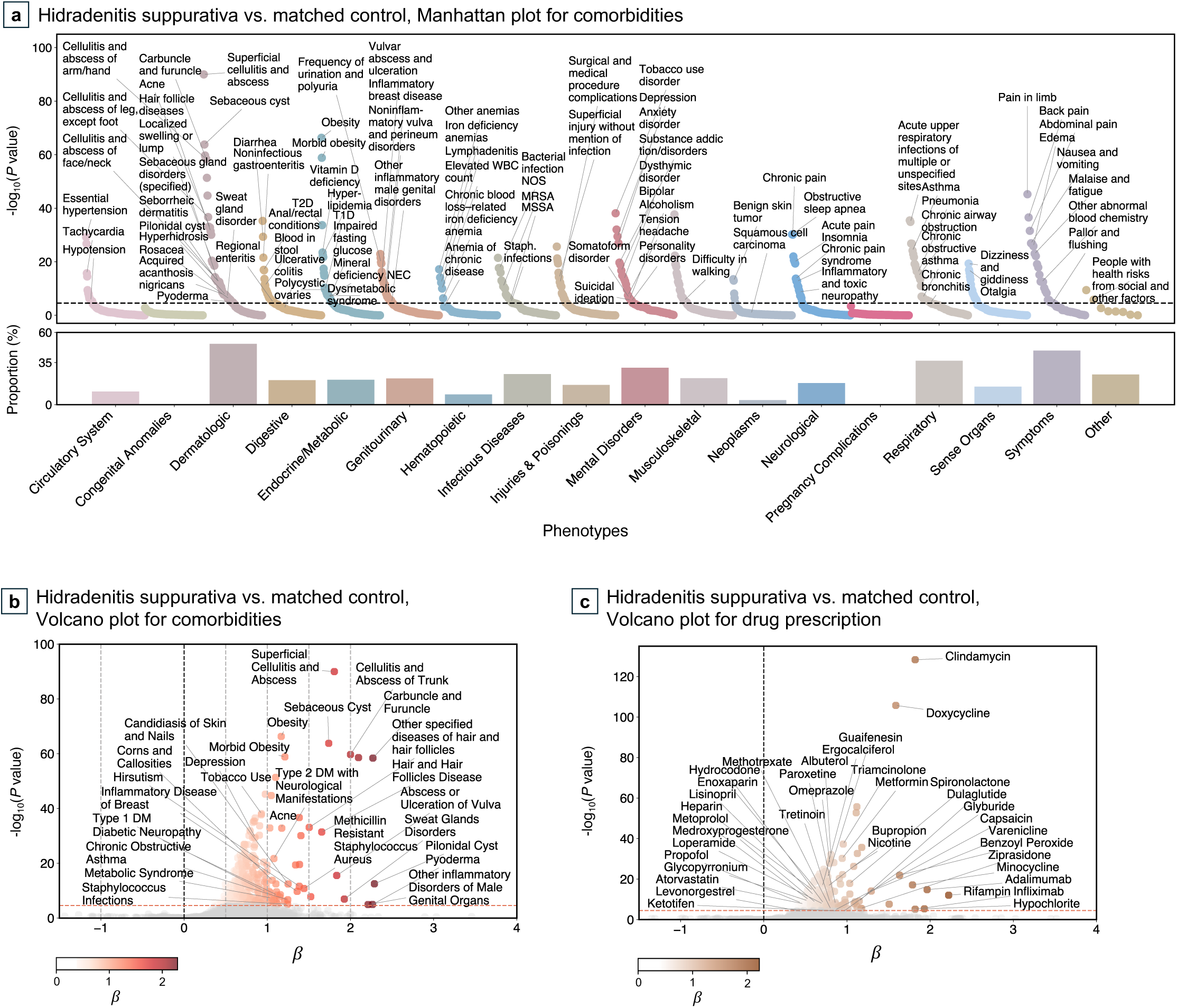
Phenotypes and medications enriched in Hidradenitis Suppurativa (HS) vs. demographic-matched controls in All of Us (Validation analysis 1). (**a**) Manhattan plot (top) showing results of comorbidity association analysis. The dots represent phecodes, colored and grouped by phecode categories. The dashed horizontal line indicates Bonferroni-corrected significance threshold. The bar plot (bottom) illustrates the proportion of phecodes within each disease category significantly enriched in HS. (**b**) Volcano plot displaying estimated coefficients of the phenotypic associations. The dashed horizontal line indicates Bonferroni-corrected significance threshold. The dots represent phecodes, colored according to their estimated association coefficients. (**c**) Volcano plot showing results of medication association analysis. The dashed horizontal line indicates Bonferroni-corrected significance threshold. The dots represent medications, colored according to their estimated association coefficients.

**Extended Data Figure 3.**
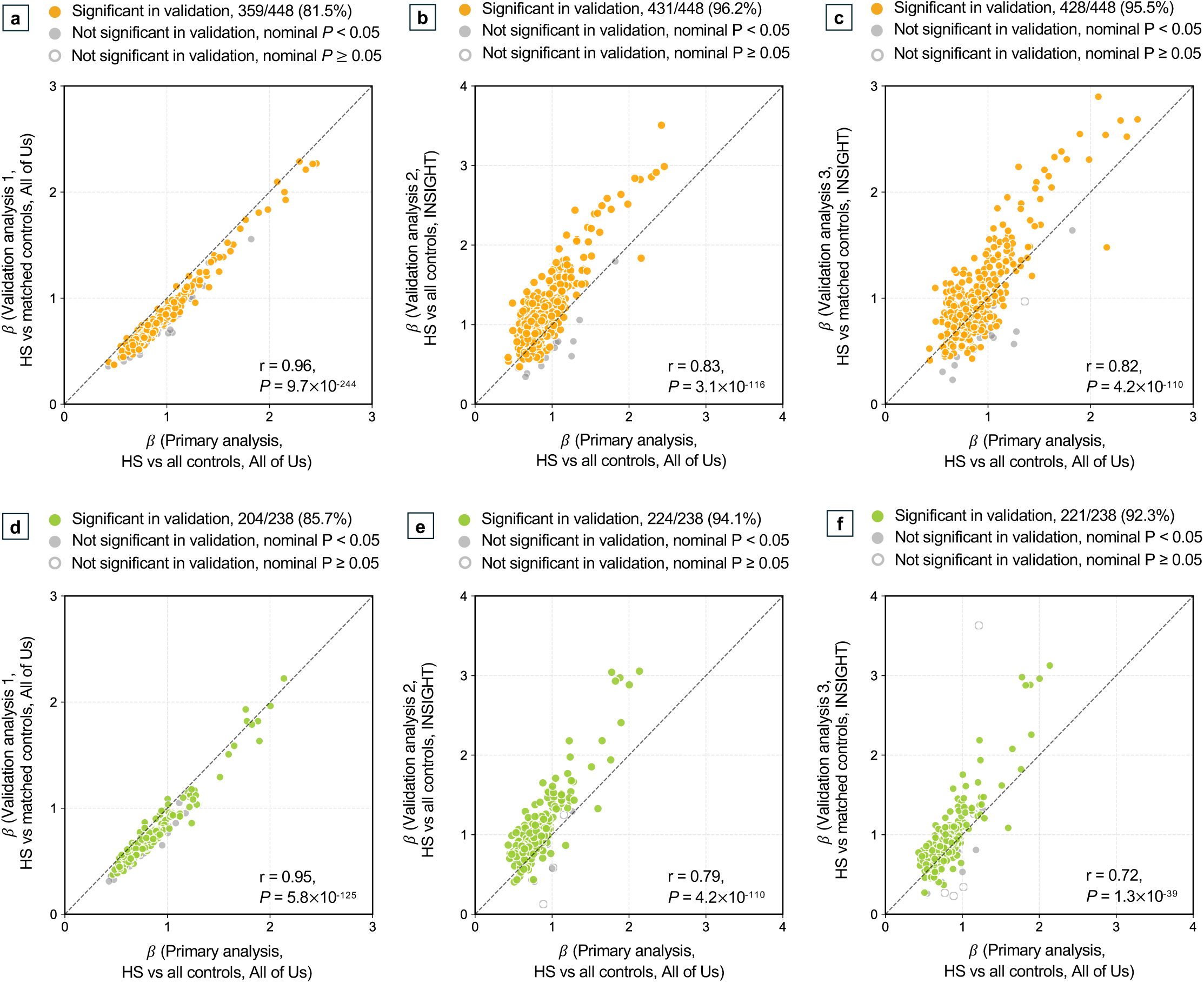
Validation of phenotypes and medications enriched in HS. We examined HS-associated phenotypes and medications detected in the primary analysis with different validation strategies: (**a**) and (**d**), HS vs. demographic-matched controls in All of Us; (**b**) and (**e**), HS vs. all controls in INSIGHT; (**c**) and (**f**), HS vs. demographic-matched controls in INSIGHT. Associations with *P* values below the Bonferroni-corrected threshold were considered significant. Pearson correlation coefficients 𝑟 quantify concordance between estimated association coefficients from the primary and validation analyses (𝑟 = 1, perfect; r = 0, none); *P* < 0.05 was considered statistically significant.

**Extended Data Figure 4.**
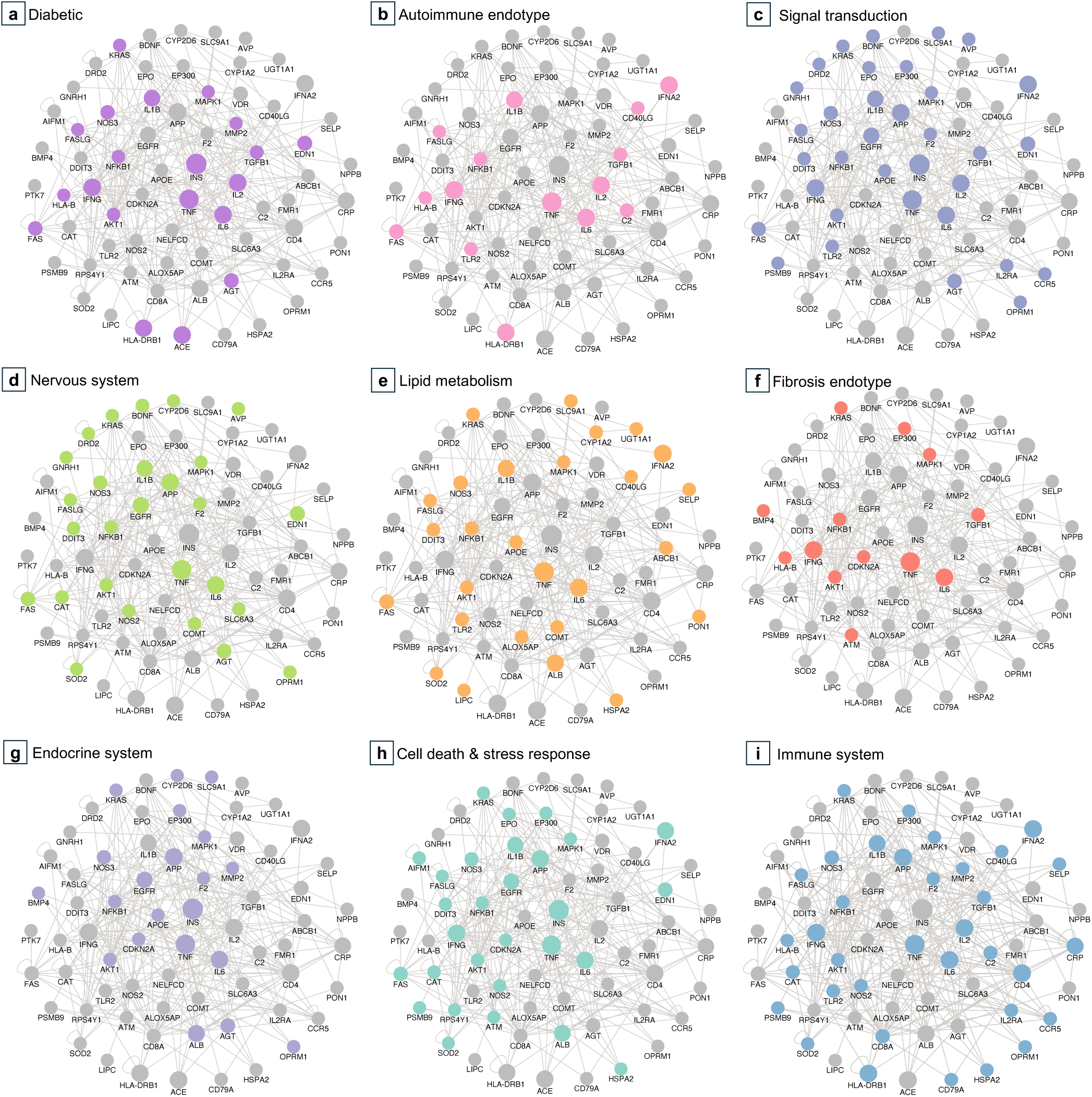
Subdivision of the phenotype-informed HS gene module (PiHSM) into nine functional endotypes. Nodes indicate PiHSM genes and edges denote known biological interactions. Node size reflects degree centrality (defined as number of connected neighbor genes in the network).

**Extended Data Figure 5.**
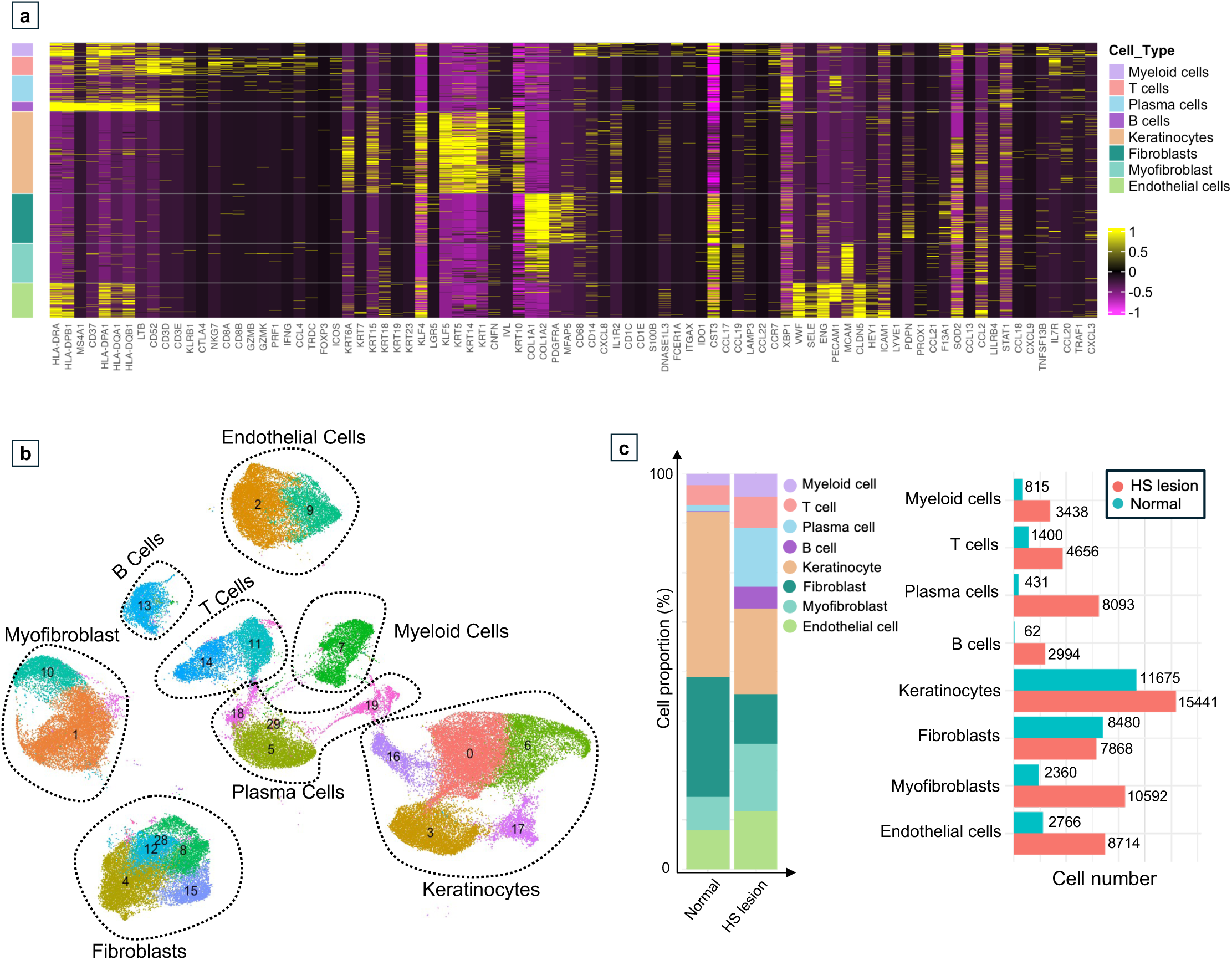
Cell type composition and marker gene expression in HS lesions and normal skin from single cell RNA sequencing. (**a**) Heatmap of canonical marker genes used to define major cell types, including myeloid cells, T cells, plasma cells, B cells, keratinocytes, fibroblasts, myofibroblasts, and endothelial cells. (**b**) UMAP visualization of cell clusters with cell type annotations. (**c**) Proportions and numbers of each cell type in HS lesion and normal samples, respectively.

**Extended Data Figure 6.**
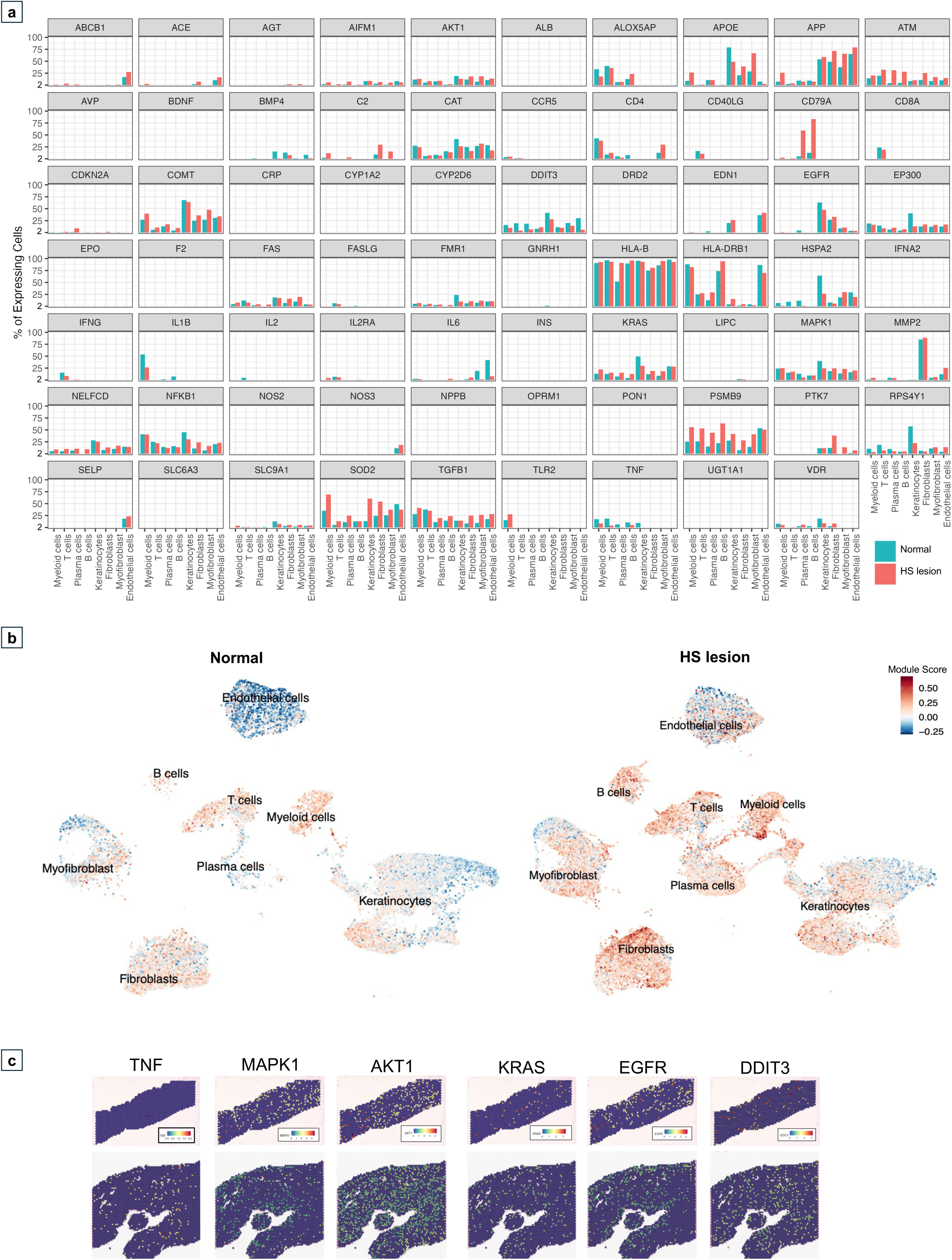
Cell type–specific gene expression and spatial localization of gene expression across conditions. (**a**) Bar plots show the percentage of cells expressing selected PiHSM genes in HS lesions and normal controls. In each subplot, the x-axis represents cell types, and the y-axis indicates the percentage of cells expressing the gene, with colors corresponding to different conditions. (**b**) UMAP plots displaying module scores, with the left panel representing cells from normal skin and the right panel representing cells from HS lesions. (**c**) Spatial transcriptomics images illustrate the spatial localization of gene expression across tissue sections.

**Extended Data Figure 7.**
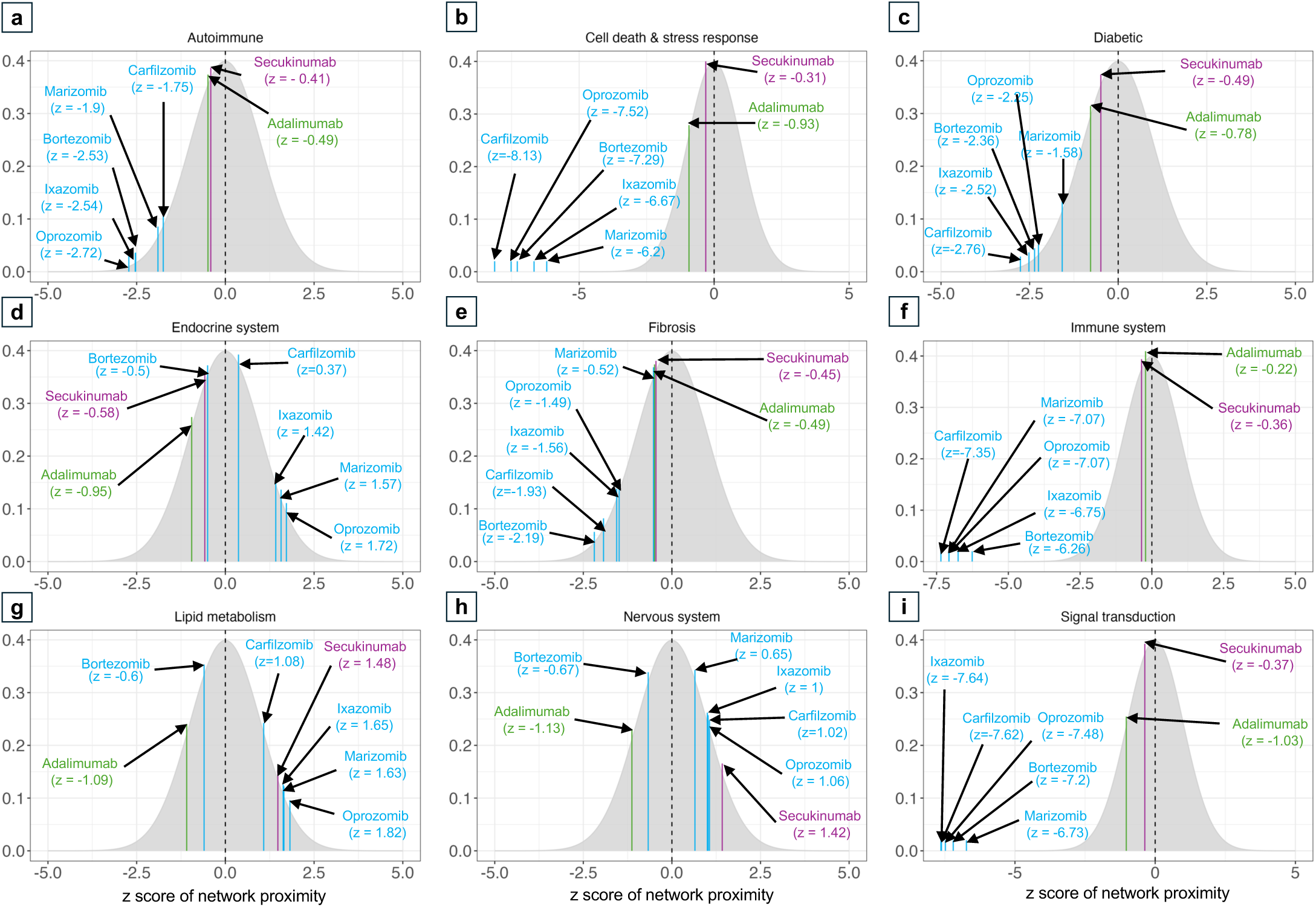
Network proximity (z-scores) of the drug candidates with functional endotype networks.

